# Application of the Sepsis-3 criteria to describe sepsis epidemiology in the Amsterdam UMCdb intensive care dataset

**DOI:** 10.1101/2023.09.24.23296037

**Authors:** Christopher YK Williams, Tom Edinburgh, Paul WG Elbers, Patrick J Thoral, Ari Ercole

## Abstract

**Introduction:** Sepsis is a major cause of morbidity and mortality worldwide. In the updated, 2016 Sepsis-3 criteria, sepsis is defined as life-threatening organ dysfunction caused by a dysregulated host response to infection, where organ dysfunction can be represented by an increase in the Sequential Organ Failure Assessment (SOFA) score of 2 points or more. We sought to apply the Sepsis-3 criteria the characterise the septic cohort in the Amsterdam University Medical Centres database (Amsterdam UMCdb).

**Methods:** We examined adult intensive care unit (ICU) admissions in the Amsterdam UMCdb, which contains de-identified data for patients admitted to a mixed surgical-medical ICU at a tertiary academic medical centre in the Netherlands. We operationalised the Sepsis-3 criteria, defining organ dysfunction as an increase in the SOFA score of 2 points or more, while infection was defined as a new course of antibiotics or an escalation in antibiotic therapy, with at least one antibiotic given intravenously. Patients with sepsis were determined to be in septic shock if they additionally required the use of vasopressors and had a lactate level >2 mmol/L.

**Results:** We identified 18,221 ICU admissions from 16,408 patients in our cohort. There were 6,371 unique sepsis episodes, of which 30.1% met the criteria for septic shock. A total of 4,958/6,371 sepsis (77.8%) episodes occurred on ICU admission. Forty-eight percent of emergency medical admissions and 37.0% of emergency surgical admissions were for sepsis. Overall, there was a 12.5% ICU mortality rate; patients with septic shock had a higher ICU mortality rate (38.5%) than those without shock (11.3%).

**Conclusions:** We successfully operationalised the Sepsis-3 criteria to the Amsterdam UMCdb, allowing the characterization and comparison of sepsis epidemiology across different centres.

## Introduction

Sepsis is a major cause of morbidity and mortality worldwide, and is associated with significant hospital costs per patient (1,2). Over the past few decades, there has been considerable focus on estimating the epidemiology of sepsis and, more recently, predicting the onset of sepsis using machine learning techniques (3–5). However, these efforts are complicated by the absence of an accepted, objective diagnostic test for sepsis, alongside previously imprecise definitions (e.g *sepsis*, *severe sepsis*, and *septicaemia*) (6).

Consequently, an expert international Sepsis-3 task force was set up in 2016, through which consensus recommendations for the definition and clinical operationalisation of sepsis were established. These recommendations were termed the *Sepsis-3* criteria, being the third iterative update following 1991 and 2001 consensus recommendations (7,8). In Sepsis-3, sepsis is defined as life-threatening organ dysfunction caused by a dysregulated host response to infection (9). To operationalise this definition, the guidelines further state that organ dysfunction can be represented by an increase in the Sequential Organ Failure Assessment (SOFA) score of 2 points or more. One proposed definition for suspected infection is the sampling of microbial cultures and administration of antibiotics within a specific time period (9). Patients with septic shock can be identified by a vasopressor requirement to maintain mean arterial pressure >65 mmHg and serum lactate levels >2 mmol/L in the absence of hypovolaemia. Crucially, the Sepsis-3 definitions were constructed to have a relationship to mortality, an essential prerequisite for face-validity.

This updated definition and recommendation for operationalising sepsis has led to studies beginning to apply the Sepsis-3 criteria to large electronic health records (EHRs) (10–12). Defining sepsis using objective criteria, rather than relying on the more subjective International Classification of Diseases (ICD) coding, is thought to provide more reliable estimates of sepsis epidemiology (13). The increasing availability of de-identified EHR datasets offers the opportunity to apply the Sepsis-3 criteria to intensive care unit (ICU) patients across a range of countries and patient cohorts, including in the United States and United Kingdom (10,12). Since ICU population, structure and practice varies throughout the world, it is important that research datasets are properly characterised. We sought to apply the Sepsis-3 criteria to characterise the septic cohort in the Amsterdam University Medical Centres database, the first freely accessible European critical care database of complete ICU records (14), in order to contribute to the existing literature evaluating sepsis epidemiology in different settings.

## Methods

### Study Population

The Amsterdam University Medical Centres database (Amsterdam UMCdb) is the first freely accessible European critical care database (14). It contains de-identified data for 20,109 patients admitted to a mixed surgical-medical intensive care unit at a tertiary academic medical centre in the Netherlands, with up to 32 critical care beds and up to 12 high-dependency beds. Patients aged 18 years and older admitted to the ICU were deemed eligible for inclusion. Exclusion criteria: patients admitted to the Medium Care Unit (MCU, high-dependency unit); patients with ICU admission duration of less than 1 hour; and patients with fewer than 3 Sequential Organ Failure Assessment (SOFA) component scores calculated on day 1 of ICU admission.

### Identification of Sepsis and Septic Shock

In this study, we operationalised the identification of sepsis using the framework described by Shah et al in characterising the Critical Care Health Informatics Collaborative UK dataset (12). Infection was defined as a new course of antibiotics or an escalation in antibiotic therapy, with at least one antibiotic given intravenously. Following the antibiotic ranking classification of Braykov et al (15), an escalation in antibiotic therapy was defined as an increase in the maximum rank of current antibiotics from one 24 hour period to the next, or an increase in the number of antibiotics with the same maximum rank prescribed (12). Organ dysfunction was defined as an increase in the Sequential Organ Failure Assessment (SOFA) score of 2 points or more (16). Patients with sepsis were determined to be in septic shock if they additionally required the use of vasopressors and had a lactate level >2 mmol/L (9).

Full details, and accompanying code, of the operationalisation of sepsis in the Amsterdam UMC database can be found here (17,18). For each patient admission in our dataset, daily SOFA scores (including both individual and total scores), daily antibiotic escalation status, and sepsis/septic shock status were calculated. Because pre-ICU data is limited in the Amsterdam UMC database, SOFA components were assumed to be zero for all patients prior to ICU admission as per the original Sepsis-3 recommendation. Patients with three or more missing SOFA components on day 0 of ICU admission were excluded. Among patients who died in the ICU, SOFA scores which could not be calculated on the day of death (due to missing SOFA components) were set to the maximum value of 24 (19). A sepsis episode was identified when the SOFA score increased by ≥ 2 on consecutive days with antibiotic escalation on either day, or when the SOFA score was at least 2 points higher on the day after antibiotic escalation compared to the day before (1).

To accurately identify clinical suspicion of infection, prophylactic administration of antibiotics must be disregarded from the calculation of antibiotic escalation status. Notably, Amsterdam UMC ICU practises selective digestive decontamination including a four-day course of cefotaxime at ICU admission. In patients with suspected infection within this time period, the hospital practice is to exchange the cefotaxime prescription for the similar-spectrum antibiotic ceftriaxone. We therefore disregarded cefotaxime prescriptions within the first four days of ICU admission when calculating antibiotic escalation status. For cefotaxime prescriptions that extended beyond the first four days of ICU admission, we assumed that a suspected infection had occurred at some point within those first four days and that the cefotaxime prescription had (erroneously) not been exchanged for ceftriaxone. Due to uncertainty over when in this time period the prescription should have been changed, when calculating sepsis in these cases we allowed the timing of a SOFA increase ≥2 to occur at any time on consecutive days between days 1 to 4 of ICU admission. Similarly, antibiotic use in the first 24 hours following ICU admission after elective surgery was assumed to be prophylactic and disregarded when calculating antibiotic escalation status. We additionally treated the following antibiotics as prophylactic (per local guidelines): vancomycin administration any day following cardiac surgery; all low dose (250 mg, 4 times daily) erythromycin administration; and all cefazoline administration.

### Statistical analysis

Mann-Whitney U and Kruskal-Wallis tests were used for statistical analysis of non-normally distributed continuous variables. The Chi-squared test was used to compare differences between categorical variables. We estimated age- and sex-adjusted hazard ratios by admission sepsis status using cause-specific Cox proportional hazard models. We carried out sensitivity analyses a) requiring >6 hours of noradrenaline infusion when calculating the cardiovascular component of SOFA score and b) where the criteria for suspected infection required both the prescription of one or more intravenous, non-prophylactic antibiotic and the presence of at least one microbial culture sample (from blood, urine, wound, catheter, faecal, drain, throat, nasal, rectal, or perineum cultures) within a 24 hour period. *p* <0.05 was considered statistically significant. Data were analysed using Python 3.9.12.

## Results

### Study characteristics

A total of 23,106 MCU/ICU admissions from the Amsterdam UMC database were examined in this study (Figure 1). Thirty-nine admissions were associated with an MCU/ICU stay duration < 1 hour and were excluded. Of the eligible admissions, 226 had fewer than three SOFA dimensions recorded in the first 24 hours and were excluded. This left 22,841 ICU/MCU admissions, of which 4,620 MCU admissions were excluded from the main analysis (Supplementary File 1, Tables S1-3). Our dataset therefore comprised 18,221 ICU admissions from 16,408 patients, including 7,397 (40.6%) elective surgical admissions, 1,741 (9.6%) emergency surgical admissions, and 9,083 (49.8%) medical admissions (Table 1; Figure S1). A majority of patients (66.0%) were over 60 years old on ICU admission, while 33.0% of patients were women. The median SOFA score was 6 (IQR 5-9) over the first 24 hours of admission. The most prevalent admitting specialty was Cardiothoracic Surgery (40.4%), followed by Cardiology (7.1%), Neurosurgery (6.0%), Gastrointestinal Surgery (5.2%), and General Internal Medicine (5.1%).

**Figure 1.**
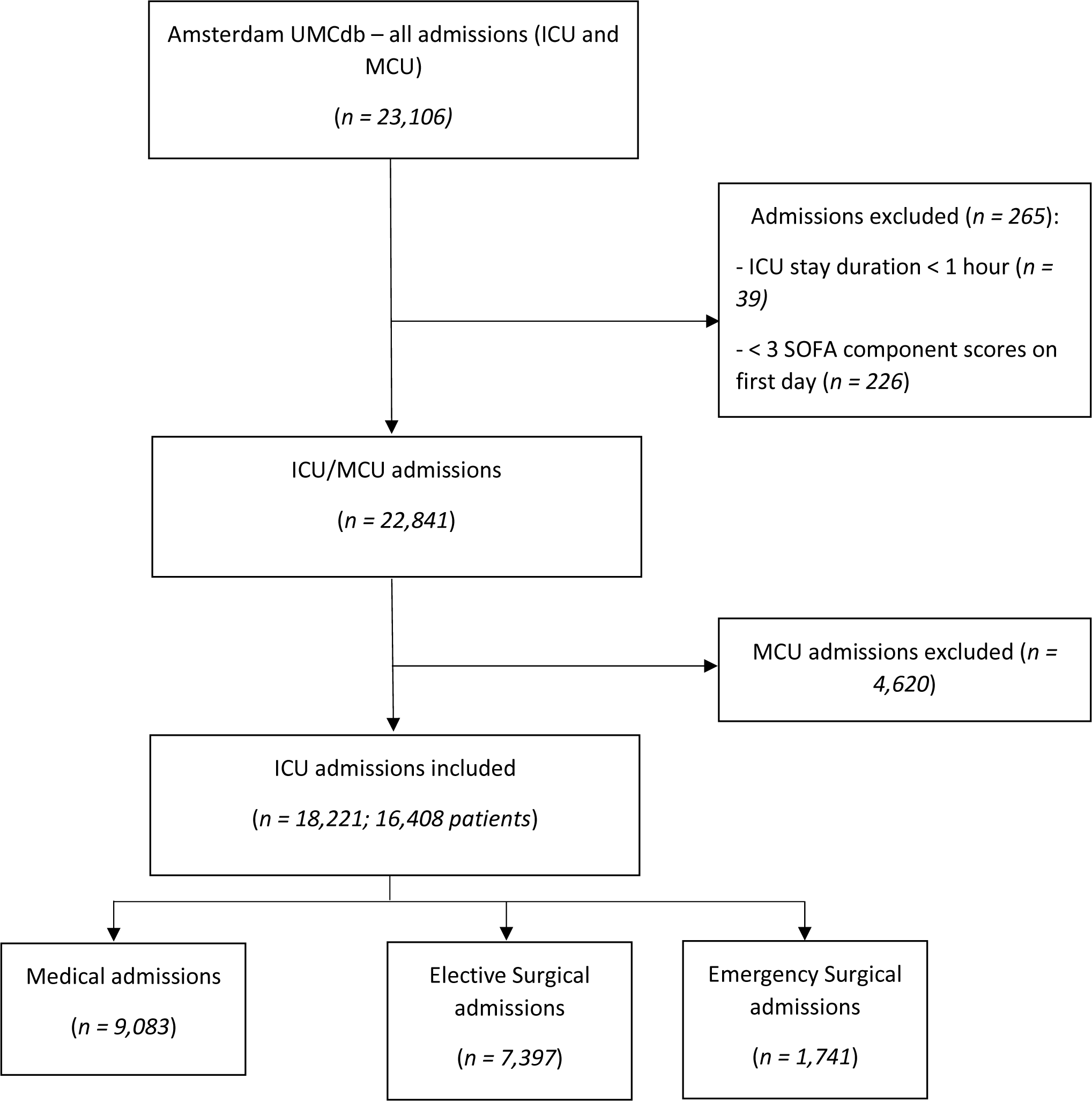
Flowchart of included ICU admissions from the Amsterdam UMC database.

**Table 1.** Summary characteristics of ICU admissions by infection status. *IV (non-prophylactic) antibiotics consecutively for the first 4 days and admission length of stay >=4 days. †IV (non-prophylactic) antibiotics for at least 4 days in total *or* until ICU death/discharge.

|  | **Overall** | ***Septic shock*** | ***Sepsis without shock*** | ***Antibiotics without sepsis*** | ***Not on antibiotics*** |
| --- | --- | --- | --- | --- | --- |
| Number of admissions | 18221 | 1654 | 3304 | 4911 | 8352 |
| Number of patients | 16408 | 1623 | 3036 | 4665 | 8135 |
| Female, n (%) | 5876 (33.0) | 607 (37.9) | 1227 (38.7) | 1744 (35.5) | 2298 (28.3) |
| **Age group, n (%)** |  |  |  |  |  |
| 18-39 | 1665 (9.1) | 203 (12.3) | 419 (12.7) | 403 (8.2) | 640 (7.7) |
| 40-49 | 1487 (8.2) | 177 (10.7) | 300 (9.1) | 421 (8.6) | 589 (7.1) |
| 50-59 | 3034 (16.7) | 247 (14.9) | 546 (16.5) | 806 (16.4) | 1435 (17.2) |
| 60-69 | 5018 (27.5) | 393 (23.8) | 830 (25.1) | 1277 (26.0) | 2518 (30.1) |
| 70-79 | 5157 (28.3) | 412 (24.9) | 860 (26.0) | 1416 (28.8) | 2469 (29.6) |
| 80+ | 1860 (10.2) | 222 (13.4) | 349 (10.6) | 588 (12.0) | 701 (8.4) |
| **Admission category, n (%)** |  |  |  |  |  |
| Elective surgical | 7397 (40.6) | 0 (0.0) | 0 (0.0) | 2866 (58.4) | 4531 (54.3) |
| Emergency surgical | 1741 (9.6) | 307 (18.6) | 337 (10.2) | 553 (11.3) | 544 (6.5) |
| Emergency medical | 9083 (49.8) | 1347 (81.4) | 2967 (89.8) | 1492 (30.4) | 3277 (39.2) |
| **Specialty, n (%)** |  |  |  |  |  |
| Cardiothoracic surgery | 7369 (40.4) | 107 (6.5) | 617 (18.7) | 1829 (37.2) | 4816 (57.7) |
| Cardiology | 1300 (7.1) | 205 (12.4) | 134 (4.1) | 735 (15.0) | 226 (2.7) |
| Neurosurgery | 1089 (6.0) | 62 (3.7) | 132 (4.0) | 457 (9.3) | 438 (5.2) |
| Gastrointestinal surgery | 942 (5.2) | 164 (9.9) | 213 (6.4) | 317 (6.5) | 248 (3.0) |
| General Internal Medicine | 921 (5.1) | 219 (13.2) | 352 (10.7) | 129 (2.6) | 221 (2.6) |
| Vascular surgery | 897 (4.9) | 73 (4.4) | 110 (3.3) | 235 (4.8) | 479 (5.7) |
| Trauma | 774 (4.2) | 109 (6.6) | 206 (6.2) | 217 (4.4) | 242 (2.9) |
| Lung disease/surgery | 637 (3.5) | 77 (4.7) | 228 (6.9) | 178 (3.6) | 154 (1.8) |
| Neurology | 568 (3.1) | 57 (3.4) | 146 (4.4) | 174 (3.5) | 191 (2.3) |
| Haematology | 233 (1.3) | 65 (3.9) | 145 (4.4) | 5 (0.1) | 18 (0.2) |
| Gynaecology | 133 (0.7) | 20 (1.2) | 38 (1.2) | 27 (0.5) | 48 (0.6) |
| Urology | 132 (0.7) | 24 (1.5) | 31 (0.9) | 49 (1.0) | 28 (0.3) |
| Other medical specialty | 1286 (7.1) | 140 (8.5) | 440 (13.3) | 88 (1.8) | 618 (7.4) |
| Other surgical specialty | 331 (1.8) | 22 (1.3) | 46 (1.4) | 120 (2.4) | 143 (1.7) |
| From other ICU | 1121 (6.2) | 232 (14.0) | 330 (10.0) | 235 (4.8) | 324 (3.9) |
| **First 24hr physiology, median (IQR)** |  |  |  |  |  |
| Maximum heart rate | 102 (89-119) | 123 (105-141) | 109 (93-127) | 101 (88-119) | 97 (87-110) |
| Minimum mean arterial pressure, mmHg | 56 (46-63) | 47 (36-55) | 56 (47-63) | 54 (43-62) | 58 (50-65) |
| Maximum FiO2 | 0.50 (0.41-0.61) | 0.70 (0.51-0.91) | 0.51 (0.41-0.80) | 0.50 (0.41-0.62) | 0.46 (0.40-0.55) |
| Minimum SpO2 | 0.94 (0.73-0.97) | 0.88 (0.72-0.95) | 0.95 (0.90-0.97) | 0.93 (0.70-0.97) | 0.95 (0.71-0.97) |
| Minimum PaO2, mmHg | 79 (68-95) | 71 (61-84) | 77 (66-94) | 78 (66-93) | 82 (71-99) |
| Minimum PaO2:FiO2 ratio | 195 (135-263) | 132 (88-198) | 180 (118-248) | 185 (128-252) | 215 (162-283) |
| Minimum GCS | 15 (10-15) | 11 (3-15) | 14 (7-15) | 15 (6-15) | 15 (14-15) |
| Maximum creatinine, μmol/L | 90 (72-119) | 132 (94-206) | 93 (71-125) | 87 (69-119) | 87 (72-106) |
| Minimum platelets | 150 (105-211) | 144 (78-216) | 163 (101-250) | 154 (108-215) | 146 (108-195) |
| Maximum bilirubin, μmol/L | 10.0 (6.0-17.0) | 14.0 (8.0-26.0) | 10.0 (6.0-17.0) | 10.0 (7.0-16.0) | 9.0 (6.0-14.0) |
| Maximum SOFA score | 6 (5-9) | 11 (8-13) | 7 (5-9) | 7 (5-10) | 5 (4-7) |
| Use of vasopressors, n (%) | 12254 (67.3) | 1654 (100.0) | 1718 (52.0) | 3688 (75.1) | 5194 (62.2) |
| Mechanical ventilation, n (%) | 15973 (87.7) | 1523 (92.1) | 2677 (81.0) | 4565 (93.0) | 7208 (86.3) |
| **Outcomes** |  |  |  |  |  |
| Antibiotic escalation, first 24hr, n (%) | 5053 (27.7) | 1654 (100.0) | 3304 (100.0) | 95 (1.9) | 0 (0.0) |
| IV antibiotics for first 4d (*), n (%) | 2453 (13.5) | 957 (57.9) | 1388 (42.0) | 71 (1.4) | 37 (0.4) |
| IV antibiotics for at least 4d (†), n (%) | 5294 (29.1) | 1389 (84.0) | 2423 (73.3) | 1101 (22.4) | 381 (4.6) |
| ICU length of stay, h, median (IQR) | 31 (22-114) | 140 (49-339) | 65 (24-201) | 62 (24-177) | 23 (20-41) |
| ICU mortality, n (%) | 2270 (12.5) | 636 (38.5) | 375 (11.3) | 808 (16.5) | 451 (5.4) |

### Identification of Sepsis and Septic Shock

Overall, we identified 33,057 occasions where SOFA score increased by ≥2 points, representing clinical deterioration, of which 6,695 (20.3%) were associated with antibiotic escalation and therefore classified as sepsis. After excluding 324 repeat sepsis classifications within 72 hours of first sepsis designation, this left 6,371 unique sepsis episodes. Among these, 2,046 (30.1%) sepsis episodes were associated with vasopressor use and lactate greater than 2 mmol/L, meeting the criteria for septic shock. There were 462 ICU admissions with >1 separate sepsis episode (434 admissions had two sepsis episodes, 28 admissions had >2 episodes), while 45 admissions had a second episode of septic shock >72 hours after the previous one.

A total of 4,958/6,371 sepsis episodes (77.8%) occurred on ICU admission, with 33.4% of these classified as septic shock on ICU admission. In a sensitivity analysis requiring a minimum of 6 hours for a noradrenaline infusion to be considered in the calculation of the cardiovascular SOFA score, approximately 150 emergency ICU admissions classified as septic shock in the initial analysis were downgraded to sepsis without shock, a difference that was statistically significant (χ2 = 9.9, *p*=0.020; Table S4, Supplementary File 1). In a second sensitivity analysis where the criteria for suspected infection required both microbial cultures to be taken and the prescription of intravenous, non-prophylactic antibiotics, there were significantly fewer emergency ICU admissions with sepsis (11.4% and 8.7% of all emergency ICU admissions had sepsis without shock and with shock, respectively, compared to 30.5% and 15.3% in the main analysis; χ2 = 2271.3, *p*<0.001; Table S5, Supplementary File 1). When stratified by admission category, 47.5% (4,314/9,083) of medical admissions were for sepsis, of which 31.2% (1,347/4,314) progressed to septic shock (Table 1). This compares to 37.0% (644/1,741) of emergency surgical admissions for sepsis, of which 47.7% (307/644) progressed to septic shock (Table 1). Seventy-three percent (2,423/3,304) of ICU admissions for sepsis without shock were associated with IV antibiotics for at least 4 days or until end of ICU stay, compared to 84.0% (1,389/1,654) admissions with septic shock.

ICU stays were longest among patients with sepsis, either without shock (median length of stay [LoS] 65 hours, interquartile range [IQR] 24-201 hours) or with septic shock (median LoS 140 hours, IQR 49-339 hours), compared to patients on antibiotics without sepsis (median LoS 62 hours, IQR 24-177 hours) and those not on antibiotics (median LoS 23 hours, IQR 20-41 hours) (*p* < 0.001). Similarly, patients who received <4 days of antibiotics (or antibiotics until ICU discharge if discharged in <4 days) had a significantly lower ICU LoS (median 34 hours, IQR 23-62 hours) compared to those receiving antibiotics for ≥4 days or until death (median 285 hours, IQR 169-518 hours) (*p*<0.001) (Table 2).

**Table 2.** Summary characteristics of ICU admissions with sepsis (with and without shock), by duration of antibiotics. *IV (non-prophylactic) antibiotics consecutively for the first 4 days and admission length of stay >=4 days. †IV (non-prophylactic) antibiotics for at least 4 days in total *or* until ICU death/discharge.

|  | **Antibiotics for at least 4 days or until death** | **Antibiotics for <4 days or until discharge (if discharged in <4 days)** | **Overall (all patients with sepsis on admission)** |
| --- | --- | --- | --- |
| Number of admissions | 2146 | 2812 | 4958 |
| Number of patients | 2005 | 2697 | 4507 |
| Women, n (%) | 783 (37.6) | 1051 (39.1) | 1834 (38.4) |
| **Age group, n (%)** |  |  |  |
| 18-39 | 283 (13.2) | 339 (12.1) | 622 (12.5) |
| 40-49 | 216 (10.1) | 261 (9.3) | 477 (9.6) |
| 50-59 | 349 (16.3) | 444 (15.8) | 793 (16.0) |
| 60-69 | 533 (24.8) | 690 (24.5) | 1223 (24.7) |
| 70-79 | 538 (25.1) | 734 (26.1) | 1272 (25.7) |
| 80+ | 227 (10.6) | 344 (12.2) | 571 (11.5) |
| **Admission category, n (%)** |  |  |  |
| Elective surgical | 0 (0.0) | 0 (0.0) | 0 (0.0) |
| Emergency surgical | 287 (13.4) | 357 (12.7) | 644 (13.0) |
| Emergency medical | 1859 (86.6) | 2455 (87.3) | 4314 (87.0) |
| **Specialty, n (%)** |  |  |  |
| Cardiothoracic surgery | 148 (6.9) | 576 (20.5) | 724 (14.6) |
| Cardiology | 159 (7.4) | 180 (6.4) | 339 (6.8) |
| Neurosurgery | 107 (5.0) | 87 (3.1) | 194 (3.9) |
| Gastrointestinal surgery | 196 (9.1) | 181 (6.4) | 377 (7.6) |
| General Internal Medicine | 293 (13.7) | 278 (9.9) | 571 (11.5) |
| Vascular surgery | 77 (3.6) | 106 (3.8) | 183 (3.7) |
| Trauma | 150 (7.0) | 165 (5.9) | 315 (6.4) |
| Lung disease/surgery | 142 (6.6) | 163 (5.8) | 305 (6.2) |
| Neurology | 95 (4.4) | 108 (3.8) | 203 (4.1) |
| Haematology | 131 (6.1) | 79 (2.8) | 210 (4.2) |
| Gynaecology | 13 (0.6) | 45 (1.6) | 58 (1.2) |
| Urology | 20 (0.9) | 35 (1.2) | 55 (1.1) |
| Other medical specialty | 206 (9.6) | 374 (13.3) | 580 (11.7) |
| Other surgical specialty | 25 (1.2) | 43 (1.5) | 68 (1.4) |
| From other ICU | 276 (12.9) | 286 (10.2) | 562 (11.3) |
| **First 24hr physiology, median (IQR)** |  |  |  |
| Maximum heart rate | 118 (101-136) | 109 (93-129) | 113 (96-132) |
| Minimum mean arterial pressure, mmHg | 48 (39-55) | 57 (47-64) | 53 (43-61) |
| Maximum FiO2 | 0.69 (0.51-0.90) | 0.50 (0.41-0.75) | 0.60 (0.41-0.90) |
| Minimum SpO2 | 0.92 (0.80-0.96) | 0.95 (0.86-0.97) | 0.94 (0.82-0.97) |
| Minimum PaO2, mmHg | 71 (62-83) | 79 (66-97) | 75 (64-91) |
| Minimum PaO2:FiO2 ratio | 131 (90-188) | 192 (127-263) | 163 (104-234) |
| Minimum GCS | 11 (3-15) | 14 (7-15) | 13 (6-15) |
| Maximum creatinine, $\mu\text{mol/L}$ | 107 (76-168) | 99 (76-141) | 102 (76-154) |
| Minimum platelets | 166 (95-255) | 149 (93-228) | 157 (94-240) |
| Maximum bilirubin, $\mu\text{mol/L}$ | 11.0 (7.0-21.0) | 11.0 (7.0-20.0) | 11.0 (7.0-20.0) |
| Maximum SOFA score | 9 (7-11) | 7 (5-9) | 8 (6-11) |
| Use of vasopressors, n (%) | 1710 (79.7) | 1662 (59.1) | 3372 (68.0) |
| Mechanical ventilation, n (%) | 1981 (92.3) | 2219 (78.9) | 4200 (84.7) |
| **Outcomes** |  |  |  |
| Antibiotic escalation, first 24hr, n (%) | 2146 (100.0) | 2812 (100.0) | 4958 (100.0) |
| IV antibiotics for first 4d (*), n (%) | 2019 (94.1) | 326 (11.6) | 2345 (47.3) |
| IV antibiotics for at least 4d (**), n (%) | 2146 (100.0) | 1666 (59.2) | 3812 (76.9) |
| ICU length of stay, h, median (IQR) | 285 (169-518) | 34 (23-62) | 85 (28-253) |
| ICU mortality, n (%) | 497 (23.2) | 514 (18.3) | 1011 (20.4) |

### Evolution of SOFA scores

Patients with septic shock on admission to the ICU had the highest maximum SOFA score on average (median 11, IQR 8-13), in contrast to patients not on antibiotics who had the lowest SOFA scores (median 5, IQR 4-7) (Table 1). Figure 2 shows component SOFA scores in the days following an ICU sepsis episode. Overall, SOFA scores improved after the start of the sepsis episode in those who survived a sepsis episode until discharge from the ICU (Figure 3) and deteriorated in those with sepsis who died in the ICU (Figure 4).

**Figure 2.**
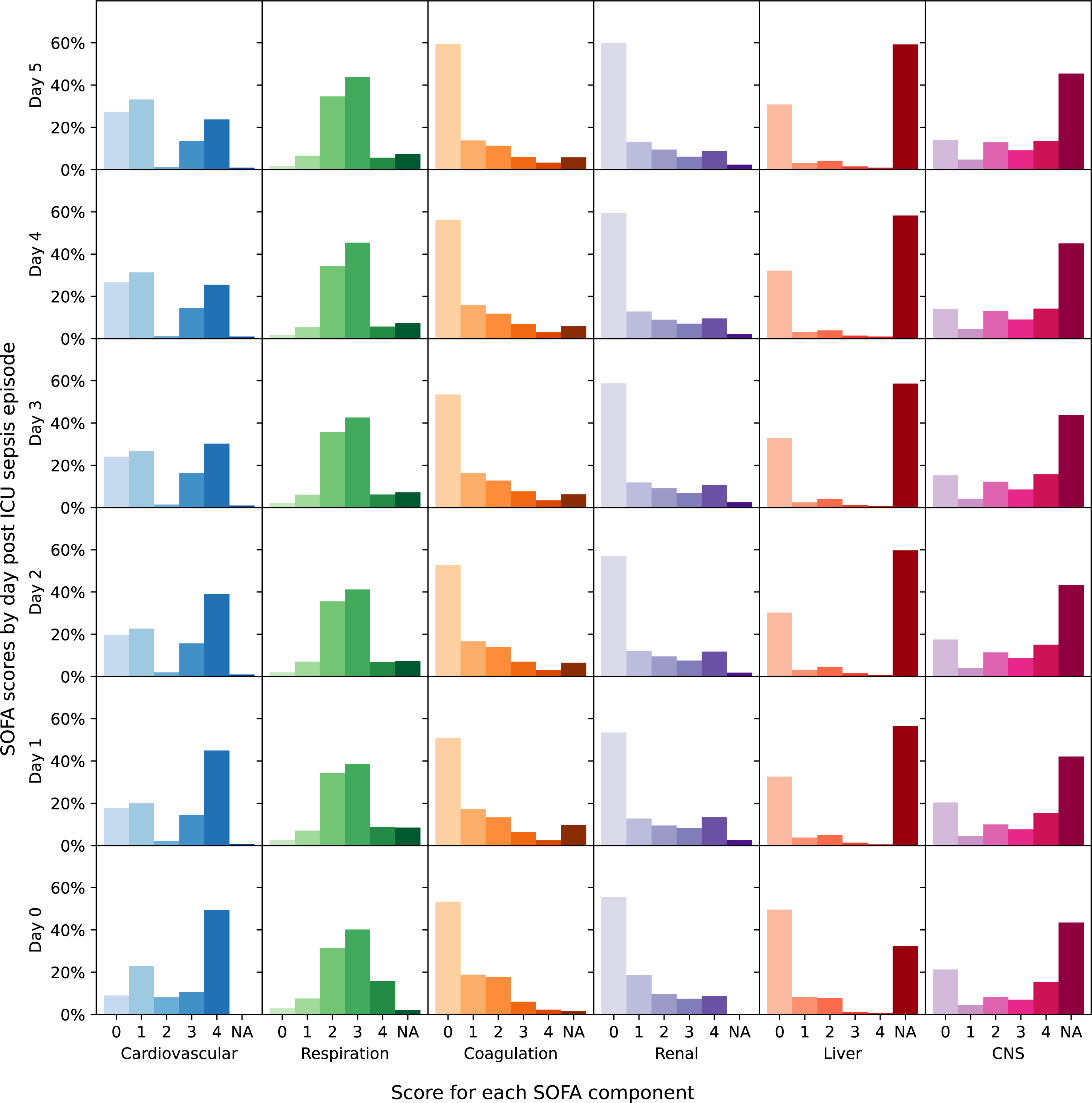
Change in component SOFA score in the days following ICU sepsis episode.

**Figure 3.**
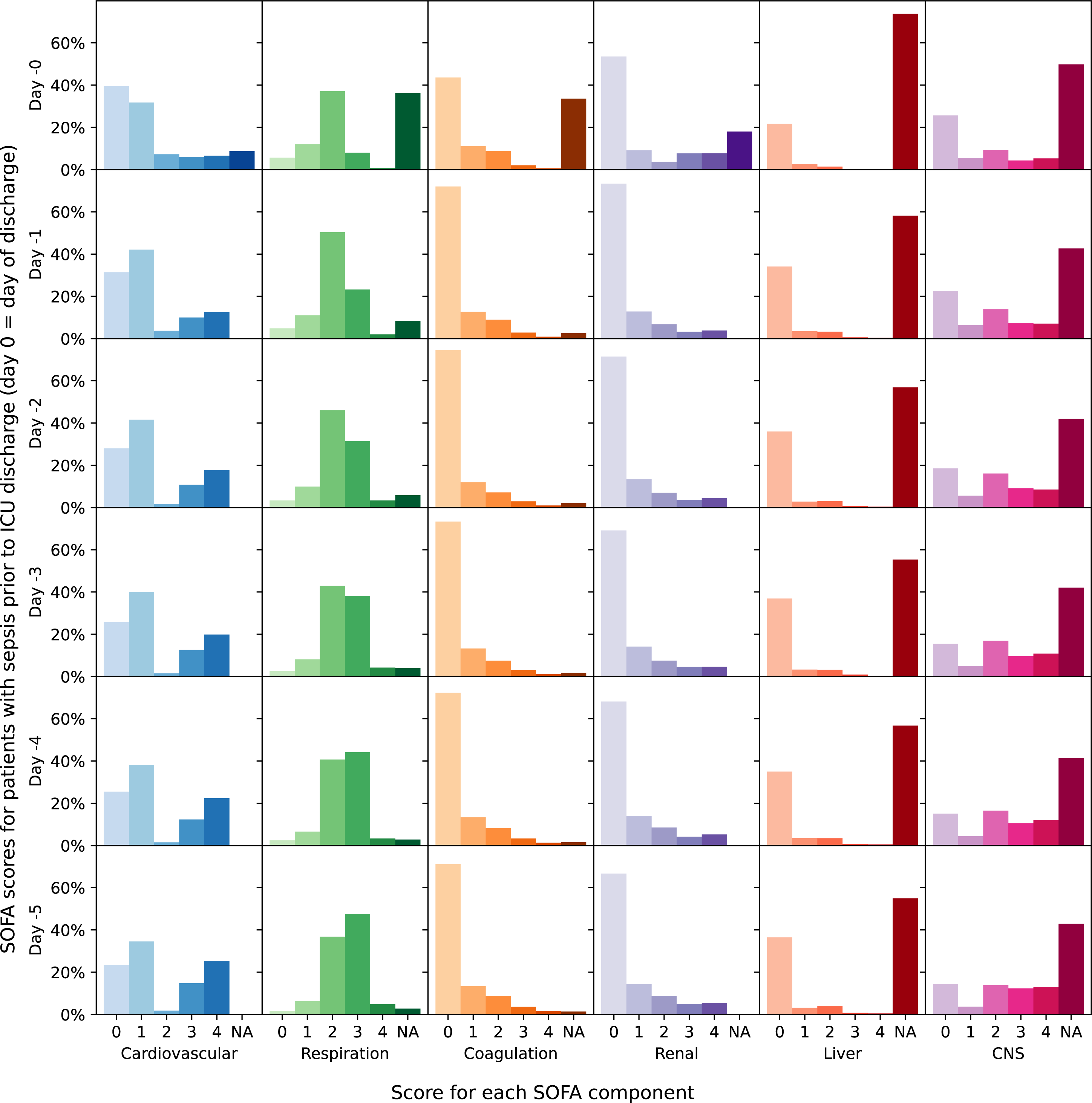
Change in component SOFA score in the 5 days prior to ICU discharge among patients with sepsis (with and without shock).

**Figure 4.**
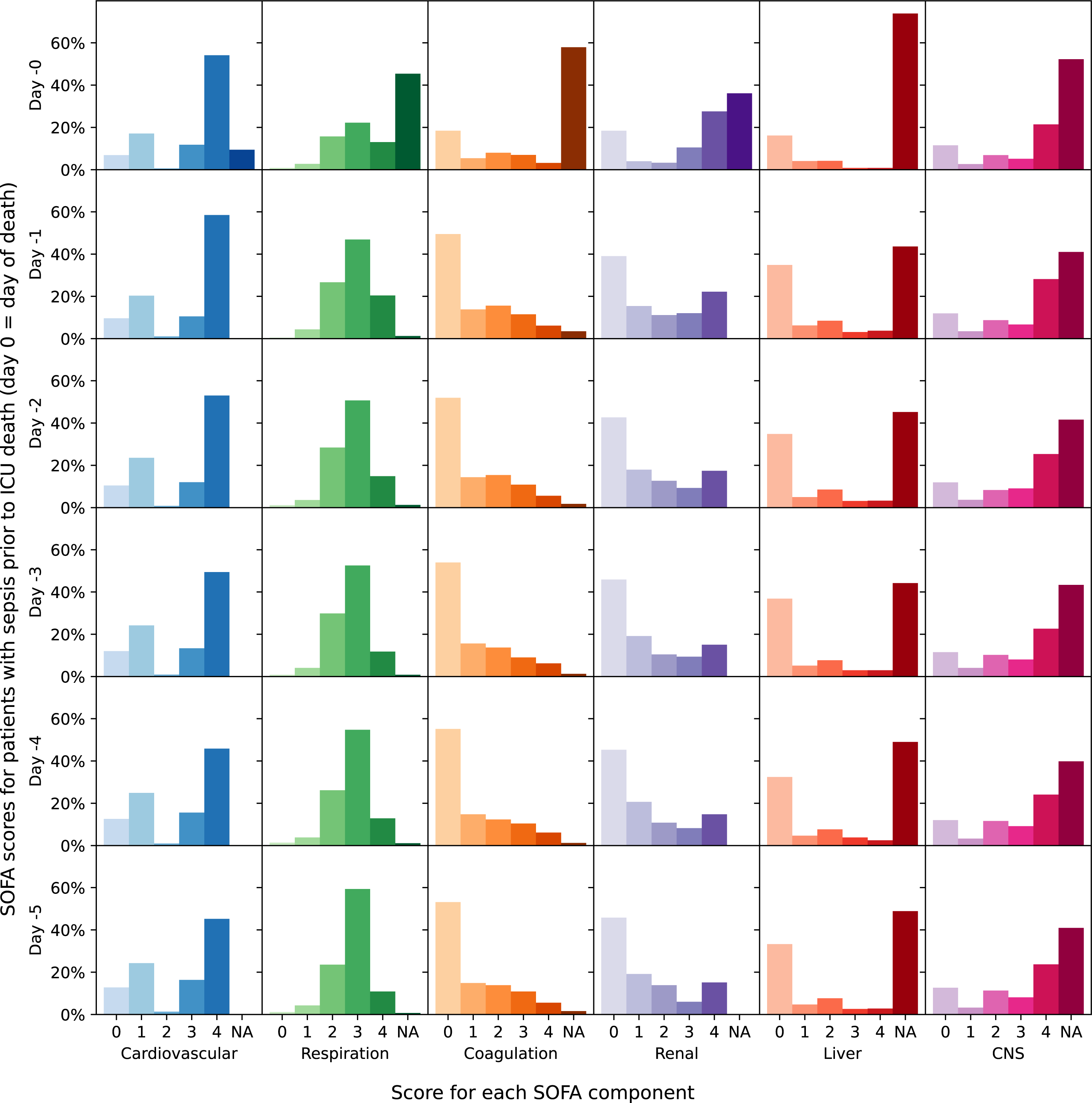
Change in component SOFA score in the 5 days prior to ICU death among patients with sepsis (with and without shock) who died intra-ICU.

### ICU mortality

Among all ICU admissions there was a 12.5% ICU mortality rate. Patients with septic shock had a higher ICU mortality rate (38.5%) than those without shock (11.3%). In addition, among all patients with sepsis (with or without shock), patients who received <4 days antibiotics had a lower mortality rate compared to those who received antibiotics for at least 4 days or until death (18.3% compared to 23.2% mortality rate, respectively) (Table 2). The trajectories of patients, stratified by sepsis status on admission, are shown in Figure 5. In age- and sex-adjusted all-mortality Cox proportional hazard models, ICU admission with septic shock and sepsis without shock was associated with hazard ratios of 1.67 (95% CI, 1.51-1.84) and 0.69 (95% CI, 0.61-0.77) respectively compared to without sepsis (Figure 6a; Table S6, Supplementary File 1). In addition, admission SOFA score was associated with a significantly increased risk of ICU mortality for all ICU admissions (Figure 6b), and for each sepsis subgroup except ‘Sepsis without shock’ (Figure 6c-f; Table S7, Supplementary File 1).

**Figure 5.**
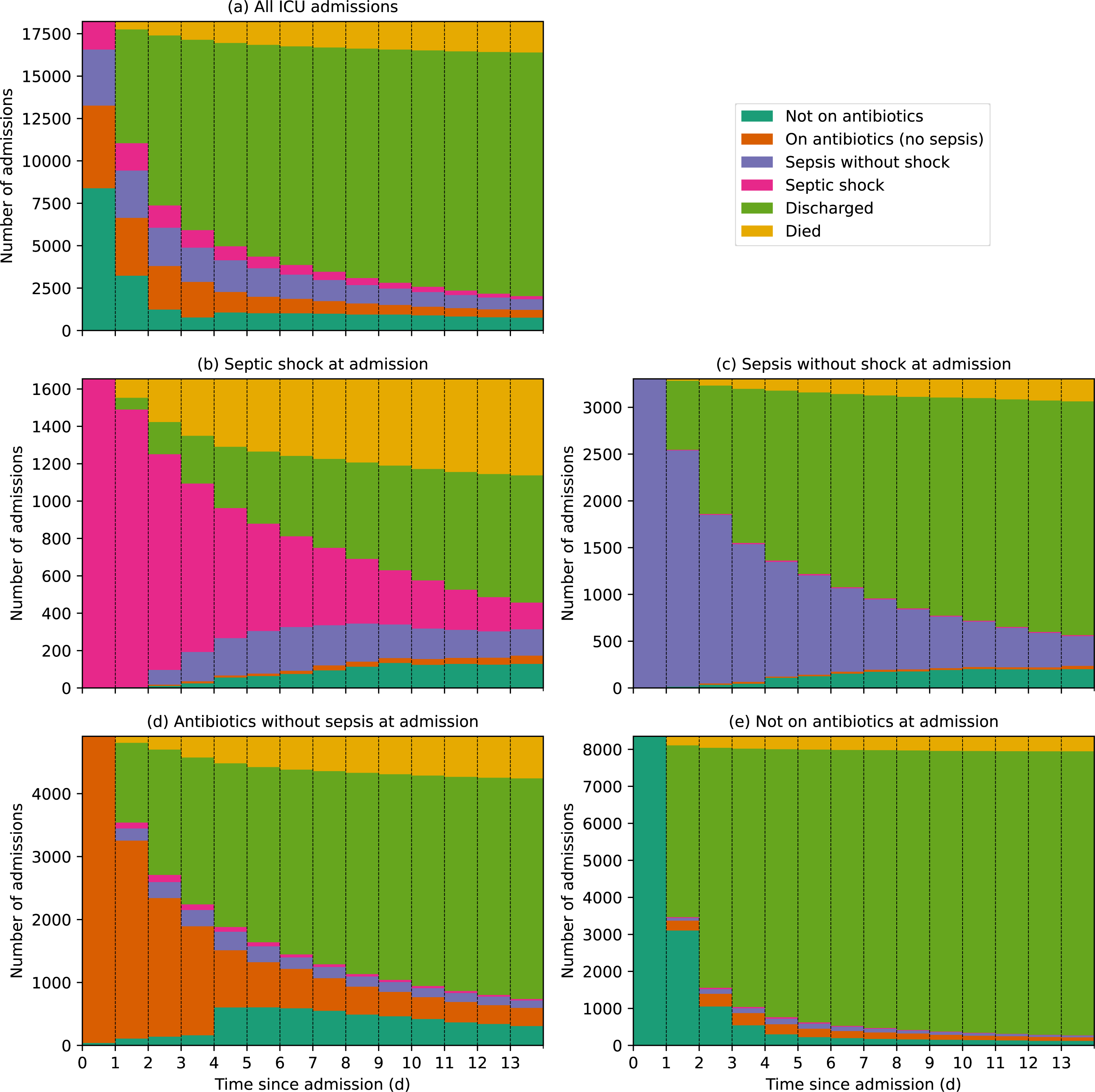
Trajectories of ICU admissions, stratified by admission sepsis status; a) All ICU admissions, b) Sepsis without shock at admission, c) Septic shock at admission, d) Antibiotics without sepsis at admission and e) Not on antibiotics at admission. We considered a patient’s admission sepsis status to remain unchanged while the following conditions were met: i) a sepsis episode continued for as long as a patient’s SOFA score is higher than their SOFA baseline* AND they remained on antibiotics, ii) a septic shock episode continued for as long as their lactate levels remained >2 or they remained on vasopressors. *Baseline SOFA score was defined as the lower of the two SOFA scores that contributed towards defining a sepsis episode; for all episodes of sepsis at ICU admission, baseline SOFA = 0.

**Figure 6.**
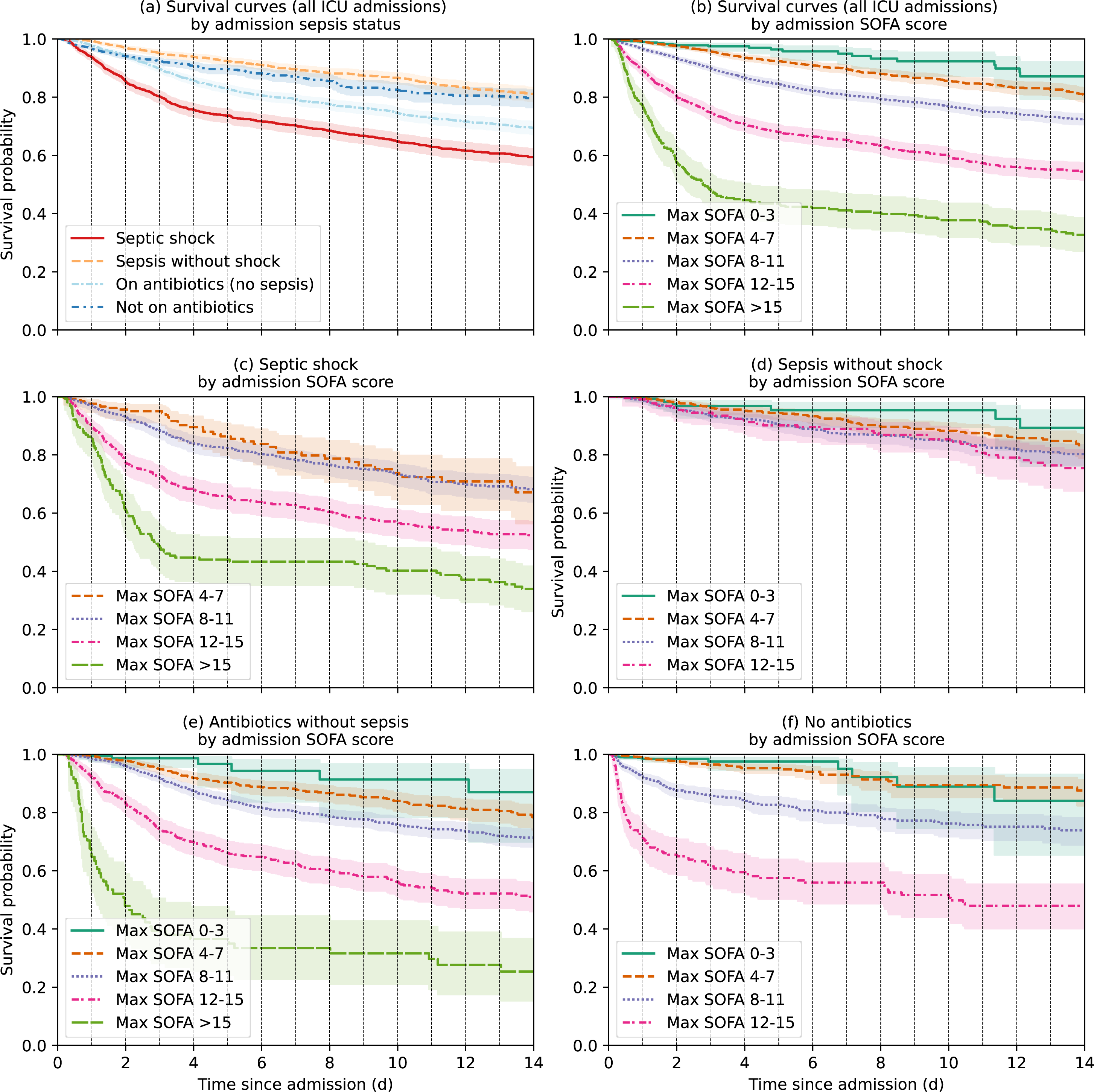
Kaplan Meier curves from cause-specific (all-mortality) Cox proportional hazard models showing survival probability based on admission sepsis status (a) and admission SOFA score (b) among all ICU admissions. 6c-f) Kaplan Meier curves showing survival probability based on admission SOFA score, among patients with (c) septic shock*, (d) sepsis without shock, (e) antibiotics without sepsis and (f) no antibiotics on ICU admission. *Due to insufficient data in the ‘*Max SOFA 0-3’* group for patients with septic shock on admission, hazard models were calculated comparing to the baseline hazard of the ‘*Max SOFA 4-7*’ group; all other hazard models in 6c-f were calculated in comparison to the baseline hazard of the ‘*Max SOFA 0-3*’ group.

### Antibiotic use

Of the 18,221 ICU admissions, 11,008 never received an antibiotic (excluding prophylactic antibiotics), 2,989 received one antibiotic regimen and 4,224 received two or more antibiotic regimens. There were 13,650 antibiotic courses prescribed over 58,341 patient-days; 21.7% of antibiotic courses were for narrow spectrum (rank 1) antibiotics, 51.6% broad spectrum (rank 2), 20.7% extended spectrum (rank 3) and 6.0% restricted (rank 4) (Table 3). The most common antibiotic prescriptions were ceftriaxone (27.1%), vancomycin (13.0%), metronidazole (9.7%), ciprofloxacin (7.7%) and co-amoxiclav (6.2%). The relative usage of treatment-only antibiotics for all patients in the ICU is shown in Figure 7.

**Figure 7.**
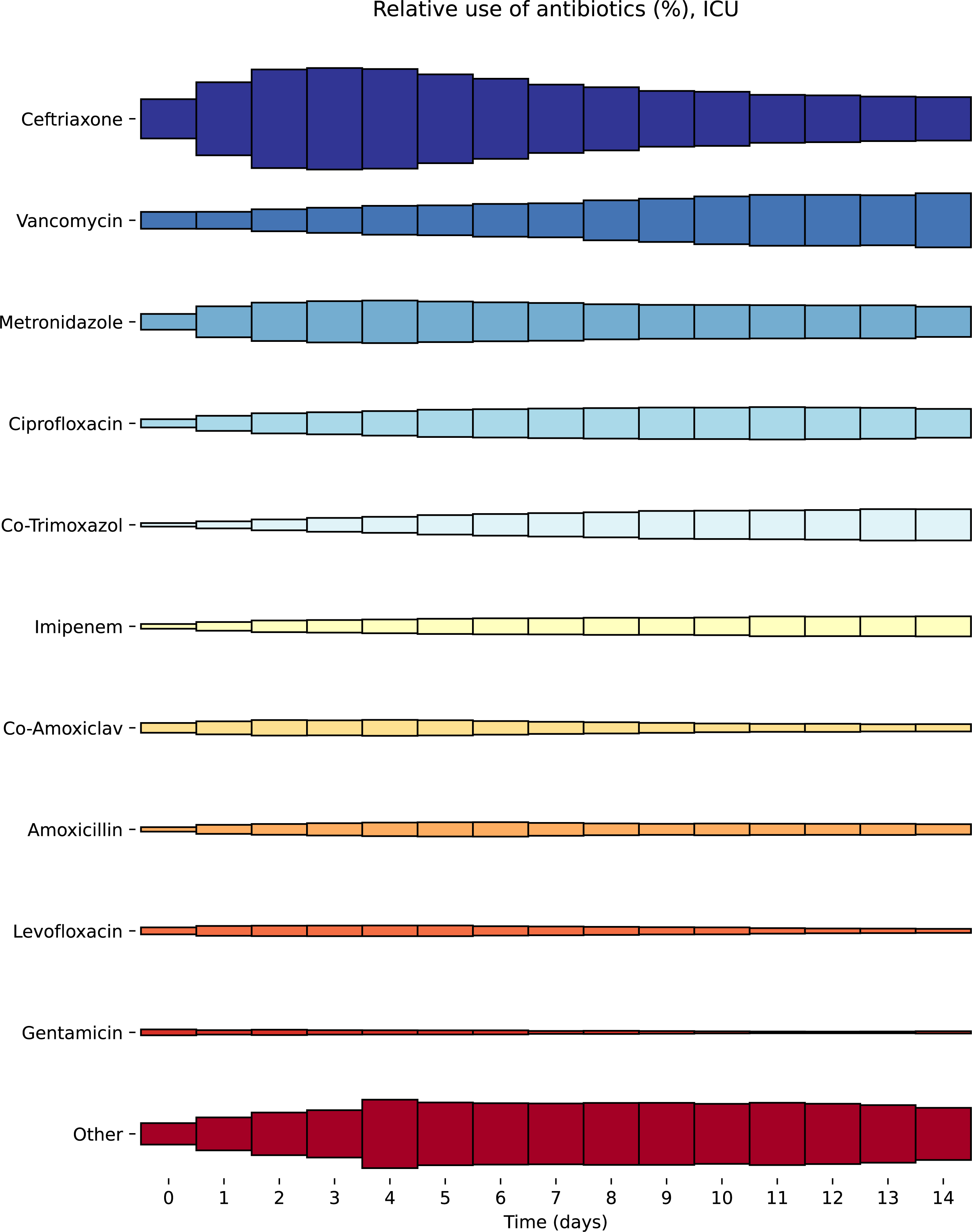
Comparison of relative antibiotic use (excluding prophylactic antibiotics) among all patients in the Amsterdam UMC intensive care unit.

**Table 3.** Antibiotic usage (treatment-only; excluding prophylactic* antibiotics) in the intensive care unit. *Prophylactic antibiotics were defined as cefotaxime prescribed within 4 days of ICU admission (as part of Amsterdam UMC’s Selective Digestive Decontamination regimen), all cefazoline administration, antibiotics prescribed within 24 hrs of admission for elective surgery patients, vancomycin administration any day following cardiac surgery, and all low dose (250 mg, 4 times daily) erythromycin administration.

| Antibiotic | Total number of courses | Percentage of all courses (%) | Total number of antibiotic-days |
| --- | --- | --- | --- |
| **Rank 4:** |  |  |  |
| Imipenem | 464 | 3.4 | 2711 |
| Meropenem | 229 | 1.7 | 1245 |
| Colistin | 108 | 0.8 | 855 |
| Amikacin | 13 | 0.1 | 55 |
| Linezolid | 4 | 0.0 | 15 |
| Neomycin sulphate | 4 | 0.0 | 12 |
| Tigecycline | 1 | 0.0 | 5 |
| Daptomycin | 0 | 0.0 | 7 |
| **Rank 3:** |  |  |  |
| Vancomycin | 1767 | 13.0 | 6381 |
| Gentamicin | 521 | 3.8 | 1014 |
| Ceftazidime | 275 | 2.0 | 1283 |
| Piperacillin | 263 | 1.9 | 1152 |
| **Rank 2:** |  |  |  |
| Ceftriaxone | 3696 | 27.1 | 15684 |
| Ciprofloxacin | 1057 | 7.7 | 4704 |
| Co-Amoxiclav | 844 | 6.2 | 2806 |
| Levofloxacin | 490 | 3.6 | 1979 |
| Erythromycin | 383 | 2.8 | 1499 |
| Cefuroxime | 295 | 2.2 | 414 |
| Clindamycin | 198 | 1.5 | 823 |
| Cefotaxime | 34 | 0.3 | 488 |
| Clarithromycin | 20 | 0.2 | 82 |
| Azithromycin | 19 | 0.1 | 52 |
| Norfloxacin | 5 | 0.0 | 7 |
| Moxifloxacin | 4 | 0.0 | 48 |
| **Rank 1:** |  |  |  |
| Metronidazole | 1326 | 9.7 | 7220 |
| Co-Trimoxazole | 627 | 4.6 | 3302 |
| Amoxicillin | 485 | 3.6 | 2263 |
| Tobramycin | 295 | 2.2 | 1100 |
| Benzylpenicillin | 174 | 1.3 | 957 |
| Doxycycline | 37 | 0.3 | 141 |
| Feneticillin | 7 | 0.1 | 23 |
| Nitrofurantoin | 5 | 0.0 | 14 |
|  | 13650 | 100.01 | 58341 |

## Discussion

### Summary of findings

In this retrospective study of 18,221 admissions to the Amsterdam University Medical Center ICU, we found that 47.5% of emergency medical admissions and 37.0% of emergency surgical admissions had sepsis, of which 31.2% and 47.7% respectively progressed to septic shock. ICU admissions with sepsis but no shock had better survival (HR 0.69 [95% CI 0.61-0.77]) than admissions without sepsis, whereas patients with septic shock had significantly worse survival (HR 1.67 [95% CI 1.51-1.84]). Higher admission SOFA scores were associated with increased ICU mortality risk, with daily SOFA scores increasing in patients who died of sepsis and decreasing among those who survived to ICU discharge.

### Comparison to other studies

By applying the Sepsis-3 criteria to Amsterdam UMCdb we were able to assess the epidemiology of sepsis in a manner that is comparable to other studies in the UK and USA (10,12). Whilst sepsis is currently defined as life-threatening organ dysfunction caused by a dysregulated host response to infection, with organ dysfunction represented by an increase in SOFA score of 2 points or more (9), there remains uncertainty between studies as to how best to define suspected infection. In a UK-based study of four National Health Service hospital trusts, Shah et al chose to define this as a new course of antibiotics or an escalation in antibiotic therapy (12). In contrast, using the publicly available MIMIC-III ICU database, Johnson et al employed the definition used by the original study from which the Sepsis-3 criteria was established, where both culture of bodily fluids and administration of antibiotics were necessary to classify infection (9,10,20).

We used the former criteria of antibiotic escalation to signify infection (regardless of culture status) in our main analysis and found comparable levels of septic shock (15.3% of emergency admissions compared to 17.9%) and overall sepsis (45.8% of emergency admissions compared to 60.4%) on ICU admission as were reported by Shah and colleagues (12). Similar to Shah et al, we also found that patients with septic shock had significantly worse survival, while those with sepsis but no shock had better survival than those with no sepsis. This latter finding may reflect the severity of non-sepsis-related diseases that warrant ICU admission. Alternatively, it is possible that, in patients admitted with sepsis but no shock, treatment is commenced sufficiently early in the disease trajectory to prevent further deterioration. Future studies should examine the effect of sepsis with and without shock on outcomes other than ICU mortality, including longer-term mortality, post-discharge co-morbidity status and overall quality of life scores.

In our sensitivity analysis requiring >6 hours of noradrenaline infusion when calculating the cardiovascular component of SOFA score, a minority of ICU admissions originally classified as having septic shock were downgraded to sepsis without shock. When the criteria for infection was modified to include both the administration of antibiotics and the sampling of bodily fluid for culture, there were significantly fewer episodes of sepsis (both with and without shock) identified: only 20.1% of emergency ICU admissions were classified as for sepsis overall (with or without shock) and 8.7% for septic shock. This compares to 49.1% of patients identified by Johnson et al who met the Sepsis-3 criteria (sepsis with or without shock) in the MIMIC-III dataset. One possible explanation for the difference in admission sepsis prevalence on using this second criteria for infection is that, due to the limited availability of pre-ICU data in the Amsterdam UMCdb, cultures may have already been taken prior to ICU admission and were consequently not repeated in-ICU at the time of antibiotic escalation. Alternatively, there may be differences in the frequency of sampling bodily fluid for culture between institutions and/or countries. It is important that this disparity between methods of determining infection status is considered when describing sepsis epidemiology across other datasets in the future.

Meanwhile, our analysis of antibiotic prescribing habits in the Amsterdam UMC intensive care unit found that ceftriaxone was the most commonly used antibiotic followed by vancomycin, metronidazole, ciprofloxacin and co-amoxiclav. Vancomycin was the most commonly used antibiotic in Rank 3 or above, with Rank 4 antibiotics rarely prescribed. Notably, Rank 3 and 4 antibiotics were used substantially less frequently (20.7% and 6.0% of antibiotic courses respectively) compared to in Shah’s study, where Rank 3 and Rank 4 antibiotics made up 31.9% and 11.1% of antibiotic courses. This may simply reflect differences in antibiotic resistance, and consequent microbiology guidance, based on location but warrants further investigation to ensure optimal antimicrobial stewardship is attained (21).

Lastly, antibiotic duration has also been suggested as one potential method of retrospectively identifying patients with true sepsis compared to those in whom antibiotics are started for suspected infection but discontinued shortly after. We found that patients admitted with sepsis who were given 4 days or more of antibiotics had greater ICU mortality (23.2%) than those in which antibiotics were discontinued within 4 days (18.3%), though our study was limited by a lack of antibiotic data for patients discharged from ICU before completing their antibiotic course. Similar ICU mortality rates have been reported elsewhere (12).

### Limitations

Our study has several limitations. Firstly, there was limited data available in the Amsterdam UMCdb for patients before and after their ICU stay. The absence of post-ICU mortality data prevented the calculation of in-hospital mortality and other longer-term post-ICU mortality rates. It is possible that the deaths of some ICU patients, in whom active treatment was withdrawn, were missed if they were transferred to the ward or other palliative settings prior to death. Similarly, the lack of data on cultures taken pre-ICU may underestimate the prevalence of suspected infection (and consequently sepsis) in our sensitivity analysis as discussed previously. Secondly, non-random missingness of data may have influenced calculation of SOFA scores and consequently alter the epidemiology of sepsis calculated. For instance, closer inspection of the daily change in component SOFA score (Figures 2-4) reveal an increasing proportion of patients where the SOFA component score could not be calculated due to missing data as time progressed. It is possible that, had SOFA scores been prospectively measured for each patient, the total SOFA score would be higher and therefore the threshold of SOFA increase ≥2 met more frequently. Finally, the ICU in this study was from an academic tertiary medical center covering a variety of specialties, including cardiothoracic surgery. This is reflected by differences in the ICU admission rate for elective surgical patients compared to other centers and consequently our results may not be representative of all ICU cohorts within the Netherlands or Europe more widely (12).

## Data Availability

All data produced are available online at https://www.amsterdammedicaldatascience.nl. It cannot be shared without going through a formal application process, because it contains detailed information of clinical care of patients, even though this information has undergone de-identification processing. Access to the data is described at https://www.amsterdammedicaldatascience.nl.

https://github.com/tedinburgh/sepsis3-amsterdamumcdb

## Acknowledgements

TE is funded by Engineering and Physical Sciences Research Council (EPSRC) National Productivity Investment Fund (NPIF) EP/S515334/1, reference 2089662. A CC BY or equivalent licence is applied to the AAM arising from this submission.

## Abbreviations

Amsterdam UMC: Amsterdam University Medical Center
EHR: Electronic health record
HR: Hazard ratio
ICD: International classification of diseases
ICU: Intensive care unit
IQR: Interquartile range
LoS: Length of stay
MCU: Medium care unit
SOFA: Sequential organ failure assessment

## Data and source code availability

The source code is available in the following repository:

- Project name: Sepsis epidemiology in Amsterdam UMCdb
- Project home page: https://github.com/tedinburgh/sepsis3-amsterdamumcdb
- Operating system(s): Platform independent
- Programming language: Python 3.9.12
- Other requirements: Python modules – numpy 1.21.5 or higher, pandas 1.3.5 or higher, re 2.2.1 or higher, argparse 1.1 or higher, lifelines 0.26.3 or higher, scipy 1.7.3 or higher, palettable 3.3.0 or higher, matplotlib 3.5.1 or higher
- License: MIT License

The dataset investigated in this article (AmsterdamUMCdb) is freely-accessible. It cannot be shared without going through a formal application process, because it contains detailed information of clinical care of patients, even though this information has undergone de-identification processing. To gain access to the database, as described at https://www.amsterdammedicaldatascience.nl, the following steps are required:

- Users must complete an appropriate training course for handling de-identified clinical data, such as Data or Specimens Only Research (DSOR) course from CITI (https://about.citiprogram.org/).
- Users must submit a signed copy of the access agreement form. Once approved, users must create an account on EASY (https://easy.dans.knaw.nl/ui/home), complete their user profile and request download permission for the dataset on EASY.

Once registered, users should then contact DANS (Data Archiving and Networked Services), who provide a link to the AmsterdamUMCdb archive.

---

**Table S1.** Summary characteristics of MCU admissions by infection status. *IV (non-prophylactic) antibiotics consecutively for the first 4 days and admission length of stay >=4 days. †IV (non-prophylactic) antibiotics for at least 4 days in total *or* until ICU death/discharge.

**Table S2.** Summary characteristics of MCU admissions with sepsis (with and without shock), stratified by duration of antibiotics. *IV (non-prophylactic) antibiotics consecutively for the first 4 days and admission length of stay >=4 days. †IV (non-prophylactic) antibiotics for at least 4 days in total *or* until ICU death/discharge.

**Table S3.** Antibiotic usage (treatment-only; excluding prophylactic* antibiotics) in the medium care unit (MCU). *Prophylactic antibiotics were defined as cefotaxime prescribed within 4 days of ICU admission (as part of Amsterdam UMC’s Selective Digestive Decontamination regimen), all cefazoline administration, antibiotics prescribed within 24 hrs of admission for elective surgery patients, vancomycin administration any day following cardiac surgery, and all low dose (250 mg, 4 times daily) erythromycin administration.

**Table S4.** Sensitivity analysis: Summary characteristics of ICU admissions by infection status when excluding noradrenaline administration <6hr in the calculation of the cardiovascular SOFA score. *IV (non-prophylactic) antibiotics consecutively for the first 4 days and admission length of stay >=4 days. †IV (non-prophylactic) antibiotics for at least 4 days in total or until ICU death/discharge.

**Table S5.** Sensitivity analysis: Summary characteristics of ICU admissions by infection status when the criteria for suspected infection required both microbial cultures to be taken and the prescription of intravenous, non-prophylactic antibiotics. *IV (non-prophylactic) antibiotics consecutively for the first 4 days and admission length of stay >=4 days. †IV (non-prophylactic) antibiotics for at least 4 days in total or until ICU death/discharge.

**Table S6.** Cause-specific Cox proportional hazard models predicting ICU mortality for *Sepsis without shock* and *Septic shock* compared to *No sepsis* among a) *all* (emergency & elective) ICU admissions and b) *emergency-only* ICU admissions, adjusted for age and sex.

**Table S7.** Cause-specific Cox proportional hazard models, adjusted for age and sex, predicting ICU mortality based on admission SOFA score (compared to baseline Max SOFA 0-3), both for all admissions (a) and grouped by sepsis status at admission (b-e). *Due to insufficient data in the ‘*Max SOFA 0-3’* group for patients with septic shock on admission, hazard models were calculated comparing to the baseline hazard of the ‘*Max SOFA 4-7*’ group; all other hazard models in S7a-S7e were calculated in comparison to the baseline hazard of the ‘*Max SOFA 0-3*’ group.

**S1 Figure.** Flowchart for (i) admission categories, (ii) sepsis at admission, (iii) number of sepsis episodes during ICU stay and (iv) outcomes. In each row/level, all boxes are subdivided (and colour coded) further by the number of admissions belonging to a combination of (i)-(iii), as shown in the Key. The size of the coloured subdivisions within each box is proportional to the number of admissions in each combined category, relative to the size of that of that box (e.g. for elective surgical patients, the subdivisions within the box for no sepsis at admission are proportional to the number of admissions who had 0, 1 and >1 sepsis episodes during ICU stay).

