## Supplementary Tables and Figure for "Application of the Sepsis-3 criteria to describe sepsis epidemiology in the Amsterdam UMCdb intensive care dataset"

**Supplementary File 1**

| | **Overall** | ***Septic shock*** | ***Sepsis without shock*** | ***Antibiotics without sepsis*** | ***Not on antibiotics*** |
|---|---|---|---|---|---|
| Number of admissions | 4620 | 63 | 589 | 551 | 3417 |
| Number of patients | 4214 | 63 | 568 | 533 | 3155 |
| Women, n (%) | 1890 (41.4) | 31 (55.4) | 223 (38.9) | 221 (40.2) | 1415 (41.8) |
| **Age group, n (%)** | | | | | |
| 18-39 | 824 (17.8) | 1 (1.6) | 77 (13.1) | 105 (19.1) | 641 (18.8) |
| 40-49 | 656 (14.2) | 8 (12.7) | 58 (9.8) | 71 (12.9) | 519 (15.2) |
| 50-59 | 874 (18.9) | 11 (17.5) | 90 (15.3) | 99 (18.0) | 674 (19.7) |
| 60-69 | 1020 (22.1) | 16 (25.4) | 149 (25.3) | 136 (24.7) | 719 (21.0) |
| 70-79 | 868 (18.8) | 19 (30.2) | 142 (24.1) | 94 (17.1) | 613 (17.9) |
| 80+ | 378 (8.2) | 8 (12.7) | 73 (12.4) | 46 (8.3) | 251 (7.3) |
| **Admission category, n (%)** | | | | | |
| Elective surgical | 2029 (43.9) | 0 (0.0) | 0 (0.0) | 354 (64.2) | 1675 (49.0) |
| Emergency surgical | 352 (7.6) | 6 (9.5) | 74 (12.6) | 35 (6.4) | 237 (6.9) |
| Emergency medical | 2239 (48.5) | 57 (90.5) | 515 (87.4) | 162 (29.4) | 1505 (44.0) |
| **Specialty, n (%)** | | | | | |
| Neurosurgery | 1424 (30.8) | 1 (1.6) | 31 (5.3) | 52 (9.4) | 1340 (39.2) |
| Vascular surgery | 534 (11.6) | 2 (3.2) | 38 (6.5) | 53 (9.6) | 441 (12.9) |
| General Internal Medicine | 418 (9.0) | 22 (34.9) | 124 (21.1) | 28 (5.1) | 244 (7.1) |
| Trauma | 379 (8.2) | 1 (1.6) | 50 (8.5) | 67 (12.2) | 261 (7.6) |
| Gastrointestinal surgery | 257 (5.6) | 8 (12.7) | 47 (8.0) | 70 (12.7) | 132 (3.9) |
| Lung disease/surgery | 217 (4.7) | 1 (1.6) | 52 (8.8) | 50 (9.1) | 114 (3.3) |
| Neurology | 209 (4.5) | 0 (0.0) | 28 (4.8) | 20 (3.6) | 161 (4.7) |
| Urology | 84 (1.8) | 1 (1.6) | 11 (1.9) | 26 (4.7) | 46 (1.3) |
| Gynaecology | 45 (1.0) | 0 (0.0) | 11 (1.9) | 11 (2.0) | 23 (0.7) |
| Cardiothoracic surgery | 34 (0.7) | 1 (1.6) | 2 (0.3) | 8 (1.5) | 23 (0.7) |
| Cardiology | 21 (0.5) | 0 (0.0) | 5 (0.8) | 1 (0.2) | 15 (0.4) |
| Haematology | 15 (0.3) | 2 (3.2) | 5 (0.8) | 3 (0.5) | 5 (0.1) |
| Other medical specialty | 356 (7.7) | 18 (28.6) | 111 (18.8) | 24 (4.4) | 203 (5.9) |
| Other surgical specialty | 143 (3.1) | 0 (0.0) | 4 (0.7) | 45 (8.2) | 94 (2.8) |
| From other ICU | 9 (0.2) | 0 (0.0) | 1 (0.2) | 2 (0.4) | 6 (0.2) |

| | | | | | |
|---|---|---|---|---|---|
| **First 24hr physiology, median (IQR)** | | | | | |
| Maximum heart rate | 100 (88-115) | 115 (102-135) | 110 (94-128) | 107 (93-124) | 98 (86-111) |
| Minimum mean arterial pressure, mmHg | 65 (54-76) | 48 (36-56) | 58 (49-67) | 62 (50-74) | 67 (57-77) |
| Maximum FiO2 | 0.27 (0.25-0.35) | 0.50 (0.35-0.90) | 0.35 (0.27-0.50) | 0.27 (0.25-0.35) | 0.27 (0.25-0.30) |
| Minimum SpO2 | 0.97 (0.95-0.98) | 0.94 (0.81-0.96) | 0.96 (0.93-0.97) | 0.97 (0.94-0.98) | 0.98 (0.96-0.99) |
| Minimum PaO2, mmHg | 95 (77-121) | 76 (68-85) | 80 (67-99) | 96 (78-122) | 100 (82-125) |
| Minimum PaO2:FiO2 ratio | 359 (254-474) | 196 (105-272) | 260 (171-352) | 364 (252-491) | 388 (294-493) |
| Minimum GCS | 15 (14-15) | 14 (11-15) | 15 (12-15) | 15 (15-15) | 15 (14-15) |
| Maximum creatinine, µmol/L | 70 (56-95) | 137 (86-256) | 89 (62-158) | 71 (56-103) | 67 (55-87) |
| Minimum platelets | 209 (159-267) | 150 (87-231) | 190 (122-261) | 226 (169-310) | 209 (164-262) |
| Maximum bilirubin, µmol/L | 9.0 (6.0-16.0) | 14.5 (7.8-28.0) | 10.0 (6.0-19.0) | 10.0 (6.0-15.0) | 9.0 (6.0-14.0) |
| Maximum SOFA score | 2.0 (1.0-4.0) | 9.0 (7.0-10.5) | 4.0 (3.0-6.0) | 2.0 (1.0-4.0) | 2.0 (1.0-4.0) |
| Use of vasopressors, n (%) | 693 (15.0) | 63 (100.0) | 137 (23.3) | 102 (18.5) | 391 (11.4) |
| Mechanical ventilation, n (%) | 288 (6.2) | 22 (34.9) | 97 (16.5) | 18 (3.3) | 151 (4.4) |
| **Outcomes** | | | | | |
| Antibiotic escalation, first 24hr, n (%) | 802 (17.4) | 63 (100.0) | 589 (100.0) | 150 (27.2) | 0 (0.0) |
| IV antibiotics for first 4d (*), n (%) | 184 (4.0) | 25 (39.7) | 135 (22.9) | 18 (3.3) | 6 (0.2) |
| IV antibiotics for at least 4d (†), n (%) | 646 (14.0) | 52 (82.5) | 410 (69.6) | 137 (24.9) | 47 (1.4) |
| ICU length of stay, h, median (IQR) | 20 (17-40) | 51 (29-117) | 38 (20-78) | 23 (18-54) | 20 (17-27) |
| ICU mortality, n (%) | 102 (2.2) | 15 (23.8) | 42 (7.1) | 9 (1.6) | 36 (1.1) |

**Table S1.** Summary characteristics of MCU admissions by infection status. *IV (non-prophylactic) antibiotics consecutively for the first 4 days and admission length of stay >=4 days. †IV (non-prophylactic) antibiotics for at least 4 days in total *or* until ICU death/discharge.

| | Antibiotics for at least 4 days or until death | Antibiotics for <4 days or until discharge (if discharged in <4 days) | Overall (all patients with sepsis on admission) |
|---|---|---|---|
| Number of admissions | 119 | 533 | 652 |
| Number of patients | 117 | 516 | 626 |
| Women, n (%) | 34 (30.4) | 220 (42.5) | 254 (40.3) |
| **Age group, n (%)** | | | |
| 18-39 | 13 (10.9) | 65 (12.2) | 78 (12.0) |
| 40-49 | 15 (12.6) | 51 (9.6) | 66 (10.1) |
| 50-59 | 12 (10.1) | 89 (16.7) | 101 (15.5) |
| 60-69 | 32 (26.9) | 133 (25.0) | 165 (25.3) |
| 70-79 | 31 (26.1) | 130 (24.4) | 161 (24.7) |
| 80+ | 16 (13.4) | 65 (12.2) | 81 (12.4) |
| **Admission category, n (%)** | | | |
| Elective surgical | 0 (0.0) | 0 (0.0) | 0 (0.0) |
| Emergency surgical | 16 (13.4) | 64 (12.0) | 80 (12.3) |
| Emergency medical | 103 (86.6) | 469 (88.0) | 572 (87.7) |
| **Specialty, n (%)** | | | |
| Neurosurgery | 6 (5.0) | 26 (4.9) | 32 (4.9) |
| Vascular surgery | 6 (5.0) | 34 (6.4) | 40 (6.1) |
| General Internal Medicine | 28 (23.5) | 118 (22.1) | 146 (22.4) |
| Trauma | 11 (9.2) | 40 (7.5) | 51 (7.8) |
| Gastrointestinal surgery | 14 (11.8) | 41 (7.7) | 55 (8.4) |
| Lung disease/surgery | 5 (4.2) | 48 (9.0) | 53 (8.1) |
| Neurology | 9 (7.6) | 19 (3.6) | 28 (4.3) |
| Urology | 1 (0.8) | 11 (2.1) | 12 (1.8) |
| Gynaecology | 0 (0.0) | 11 (2.1) | 11 (1.7) |
| Cardiothoracic surgery | 2 (1.7) | 1 (0.2) | 3 (0.5) |
| Cardiology | 0 (0.0) | 5 (0.9) | 5 (0.8) |
| Haematology | 1 (0.8) | 6 (1.1) | 7 (1.1) |
| Other medical specialty | 32 (26.9) | 97 (18.2) | 129 (19.8) |
| Other surgical specialty | 0 (0.0) | 4 (0.8) | 4 (0.6) |
| From other ICU | 0 (0.0) | 1 (0.2) | 1 (0.2) |

| | | | |
|---|---|---|---|
| **First 24hr physiology, median (IQR)** | | | |
| Maximum heart rate | 123 (106-140) | 107 (94-125) | 110 (95-129) |
| Minimum mean arterial pressure, mmHg | 46 (36-56) | 59 (51-68) | 57 (48-66) |
| Maximum FiO2 | 0.40 (0.30-0.60) | 0.35 (0.27-0.50) | 0.35 (0.27-0.50) |
| Minimum SpO2 | 0.95 (0.91-0.96) | 0.96 (0.92-0.98) | 0.96 (0.92-0.97) |
| Minimum PaO2, mmHg | 74 (62-83) | 81 (70-99) | 79 (68-97) |
| Minimum PaO2:FiO2 ratio | 200 (131-300) | 263 (178-351) | 255 (161-343) |
| Minimum GCS | 15 (10-15) | 15 (13-15) | 15 (12-15) |
| Maximum creatinine, µmol/L | 89 (62-146) | 93 (65-167) | 93 (64-164) |
| Minimum platelets | 218 (111-298) | 184 (117-255) | 186 (117-259) |
| Maximum bilirubin, µmol/L | 13.0 (7.0-25.0) | 10.0 (6.0-19.0) | 11.0 (6.0-21.0) |
| Maximum SOFA score | 6 (4-8) | 4 (3-6) | 4 (3-7) |
| Use of vasopressors, n (%) | 44 (37.0) | 156 (29.3) | 200 (30.7) |
| Mechanical ventilation, n (%) | 38 (31.9) | 81 (15.2) | 119 (18.3) |
| **Outcomes** | | | |
| Antibiotic escalation, first 24hr, n (%) | 119 (100.0) | 533 (100.0) | 652 (100.0) |
| IV antibiotics for first 4d (*), n (%) | 110 (92.4) | 50 (9.4) | 160 (24.5) |
| IV antibiotics for at least 4d (**), n (%) | 119 (100.0) | 343 (64.4) | 462 (70.9) |
| ICU length of stay, h, median (IQR) | 201 (130-334) | 29 (19-51) | 39 (21-80) |
| ICU mortality, n (%) | 18 (15.1) | 39 (7.3) | 57 (8.7) |

**Table S2.** Summary characteristics of MCU admissions with sepsis (with and without shock), stratified by duration of antibiotics. *IV (non-prophylactic) antibiotics consecutively for the first 4 days and admission length of stay >=4 days. †IV (non-prophylactic) antibiotics for at least 4 days in total *or* until ICU death/discharge.

| Antibiotic | Total number of courses | Percentage of all courses (%) | Total number of antibiotic-days |
|---|---|---|---|
| **Rank 4:** | | | |
| Imipenem | 40 | 2.55 | 85 |
| Meropenem | 22 | 1.4 | 101 |
| Colistin | 18 | 1.15 | 149 |
| Linezolid | 2 | 0.13 | 7 |
| Amikacin | 1 | 0.06 | 13 |
| Aztreonam | 1 | 0.06 | 1 |
| **Rank 3:** | | | |
| Vancomycin | 119 | 7.59 | 358 |
| Gentamicin | 40 | 2.55 | 94 |
| Ceftazidime | 39 | 2.49 | 112 |
| Piperacillin | 12 | 0.77 | 61 |
| **Rank 2:** | | | |
| Ceftriaxone | 433 | 27.61 | 1150 |
| Cefuroxime | 158 | 10.08 | 224 |
| Co-Amoxiclav | 148 | 9.44 | 420 |
| Ciprofloxacin | 98 | 6.25 | 352 |
| Levofloxacin | 35 | 2.23 | 81 |
| Clindamycin | 34 | 2.17 | 86 |
| Erythromycin | 23 | 1.47 | 54 |
| Azithromycin | 10 | 0.64 | 14 |
| Cefotaxime | 2 | 0.13 | 35 |
| Clarithromycin | 2 | 0.13 | 4 |
| Moxifloxacin | 2 | 0.13 | 7 |
| **Rank 1:** | | | |
| Metronidazole | 185 | 11.8 | 583 |
| Amoxicillin | 55 | 3.51 | 161 |
| Co-Trimoxazole | 50 | 3.19 | 174 |
| Tobramycin | 18 | 1.15 | 61 |
| Benzylpenicillin | 12 | 0.77 | 40 |
| Nitrofurantoin | 7 | 0.45 | 22 |
| Doxycycline | 2 | 0.13 | 9 |

| | | | |
|---|---|---|---|
| | 1568 | 100 | 4458 |

**Table S3.** Antibiotic usage (treatment-only; excluding prophylactic* antibiotics) in the medium care unit (MCU). *Prophylactic antibiotics were defined as cefotaxime prescribed within 4 days of ICU admission (as part of Amsterdam UMC's Selective Digestive Decontamination regimen), all cefazoline administration, antibiotics prescribed within 24 hrs of admission for elective surgery patients, vancomycin administration any day following cardiac surgery, and all low dose (250 mg, 4 times daily) erythromycin administration.

| | **Overall** | ***Septic shock*** | ***Sepsis without shock*** | ***Antibiotics without sepsis*** | ***Not on antibiotics*** |
|---|---|---|---|---|---|
| Number of admissions | 18221 | 1507 | 3447 | 4915 | 8352 |
| Number of patients | 16408 | 1482 | 3171 | 4668 | 8135 |
| Women, n (%) | 5876 (33.0) | 545 (37.2) | 1286 (38.9) | 1747 (35.6) | 2298 (28.3) |
| **Age group, n (%)** | | | | | |
| 18-39 | 1665 (9.1) | 179 (11.9) | 441 (12.8) | 405 (8.2) | 640 (7.7) |
| 40-49 | 1487 (8.2) | 167 (11.1) | 310 (9.0) | 421 (8.6) | 589 (7.1) |
| 50-59 | 3034 (16.7) | 222 (14.7) | 571 (16.6) | 806 (16.4) | 1435 (17.2) |
| 60-69 | 5018 (27.5) | 360 (23.9) | 861 (25.0) | 1279 (26.0) | 2518 (30.1) |
| 70-79 | 5157 (28.3) | 377 (25.0) | 895 (26.0) | 1416 (28.8) | 2469 (29.6) |
| 80+ | 1860 (10.2) | 202 (13.4) | 369 (10.7) | 588 (12.0) | 701 (8.4) |
| **Admission category, n (%)** | | | | | |
| Elective surgical | 7397 (40.6) | 0 (0.0) | 0 (0.0) | 2866 (58.3) | 4531 (54.3) |
| Emergency surgical | 1741 (9.6) | 275 (18.2) | 367 (10.6) | 555 (11.3) | 544 (6.5) |
| Emergency medical | 9083 (49.8) | 1232 (81.8) | 3080 (89.4) | 1494 (30.4) | 3277 (39.2) |
| **Specialty, n (%)** | | | | | |
| Cardiothoracic surgery | 7369 (40.4) | 73 (4.8) | 651 (18.9) | 1829 (37.2) | 4816 (57.7) |
| Cardiology | 1300 (7.1) | 195 (12.9) | 144 (4.2) | 735 (15.0) | 226 (2.7) |
| Neurosurgery | 1089 (6.0) | 60 (4.0) | 133 (3.9) | 458 (9.3) | 438 (5.2) |
| Gastrointestinal surgery | 942 (5.2) | 160 (10.6) | 216 (6.3) | 318 (6.5) | 248 (3.0) |
| General Internal Medicine | 921 (5.1) | 208 (13.8) | 363 (10.5) | 129 (2.6) | 221 (2.6) |
| Vascular surgery | 897 (4.9) | 64 (4.2) | 118 (3.4) | 236 (4.8) | 479 (5.7) |
| Trauma | 774 (4.2) | 101 (6.7) | 213 (6.2) | 218 (4.4) | 242 (2.9) |
| Lung disease/surgery | 637 (3.5) | 68 (4.5) | 237 (6.9) | 178 (3.6) | 154 (1.8) |
| Neurology | 568 (3.1) | 51 (3.4) | 152 (4.4) | 174 (3.5) | 191 (2.3) |
| Haematology | 233 (1.3) | 63 (4.2) | 147 (4.3) | 5 (0.1) | 18 (0.2) |
| Gynaecology | 133 (0.7) | 17 (1.1) | 41 (1.2) | 27 (0.5) | 48 (0.6) |
| Urology | 132 (0.7) | 22 (1.5) | 33 (1.0) | 49 (1.0) | 28 (0.3) |
| Other medical specialty | 1286 (7.1) | 126 (8.4) | 454 (13.2) | 88 (1.8) | 618 (7.4) |
| Other surgical specialty | 331 (1.8) | 17 (1.1) | 51 (1.5) | 120 (2.4) | 143 (1.7) |
| From other ICU | 1121 (6.2) | 211 (14.0) | 351 (10.2) | 235 (4.8) | 324 (3.9) |

| | | | | | |
|---|---|---|---|---|---|
| **First 24hr physiology, median (IQR)** | | | | | |
| Maximum heart rate | 102 (89-119) | 124 (106-142) | 109 (93-128) | 101 (88-119) | 97 (87-110) |
| Minimum mean arterial pressure, mmHg | 56 (46-63) | 46 (36-54) | 55 (47-63) | 54 (43-62) | 58 (50-65) |
| Maximum FiO2 | 0.50 (0.41-0.61) | 0.71 (0.51-0.98) | 0.51 (0.41-0.80) | 0.50 (0.41-0.62) | 0.46 (0.40-0.55) |
| Minimum SpO2 | 0.94 (0.73-0.97) | 0.87 (0.72-0.95) | 0.95 (0.90-0.97) | 0.93 (0.70-0.97) | 0.95 (0.71-0.97) |
| Minimum PaO2, mmHg | 79 (68-95) | 70 (60-83) | 77 (66-94) | 78 (66-93) | 82 (71-99) |
| Minimum PaO2:FiO2 ratio | 195 (135-263) | 127 (86-189) | 180 (118-249) | 185 (128-253) | 215 (162-283) |
| Minimum GCS | 15 (10-15) | 11.0 (3.0-15.0) | 14.0 (7.0-15.0) | 15.0 (6.0-15.0) | 15 (14-15) |
| Maximum creatinine, µmol/L | 90 (72-119) | 136 (95-210) | 93 (72-126) | 87 (69-119) | 87 (72-106) |
| Minimum platelets | 150 (105-211) | 146 (78-219) | 161 (100-248) | 154 (108-215) | 146 (108-195) |
| Maximum bilirubin, µmol/L | 10.0 (6.0-17.0) | 14.0 (8.0-26.0) | 10.0 (6.0-17.0) | 10.0 (7.0-16.0) | 9.0 (6.0-14.0) |
| Maximum SOFA score | 6.0 (5.0-9.0) | 11.0 (9.0-13.0) | 7.0 (5.0-9.0) | 7.0 (5.0-10.0) | 5.0 (4.0-7.0) |
| Use of vasopressors, n (%) | 12254 (67.3) | 1507 (100.0) | 1861 (54.0) | 3692 (75.1) | 5194 (62.2) |
| Mechanical ventilation, n (%) | 15973 (87.7) | 1395 (92.6) | 2803 (81.3) | 4567 (92.9) | 7208 (86.3) |
| **Outcomes** | | | | | |
| Antibiotic escalation, first 24hr, n (%) | 5053 (27.7) | 1507 (100.0) | 3447 (100.0) | 99 (2.0) | 0 (0.0) |
| IV antibiotics for first 4d (*), n (%) | 2453 (13.5) | 911 (60.5) | 1434 (41.6) | 71 (1.4) | 37 (0.4) |
| IV antibiotics for at least 4d (**), n (%) | 5294 (29.1) | 1276 (84.7) | 2536 (73.6) | 1101 (22.4) | 381 (4.6) |
| ICU length of stay, h, median (IQR) | 31 (22-114) | 160 (57-361) | 64 (24-196) | 62 (24-177) | 23 (20-41) |
| ICU mortality, n (%) | 2270 (12.5) | 609 (40.4) | 401 (11.6) | 809 (16.5) | 451 (5.4) |

**Table S4.** Sensitivity analysis: Summary characteristics of ICU admissions by infection status when excluding noradrenaline administration <6hr in the calculation of the cardiovascular SOFA score. *IV (non-prophylactic) antibiotics consecutively for the first 4 days and admission length of stay >=4 days. †IV (non-prophylactic) antibiotics for at least 4 days in total or until ICU death/discharge.

| | **Overall** | ***Septic shock*** | ***Sepsis without shock*** | ***Antibiotics without sepsis*** | ***Not on antibiotics*** |
|---|---|---|---|---|---|
| Number of admissions | 18221 | 939 | 1233 | 7697 | 8352 |
| Number of patients | 16408 | 924 | 1162 | 7162 | 8135 |
| Women, n (%) | 5876 (33.0) | 351 (38.1) | 459 (38.4) | 2768 (36.6) | 2298 (28.3) |
| **Age group, n (%)** | | | | | |
| 18-39 | 1665 (9.1) | 104 (11.1) | 161 (13.1) | 760 (9.9) | 640 (7.7) |
| 40-49 | 1487 (8.2) | 107 (11.4) | 112 (9.1) | 679 (8.8) | 589 (7.1) |
| 50-59 | 3034 (16.7) | 145 (15.4) | 212 (17.2) | 1242 (16.1) | 1435 (17.2) |
| 60-69 | 5018 (27.5) | 224 (23.9) | 312 (25.3) | 1964 (25.5) | 2518 (30.1) |
| 70-79 | 5157 (28.3) | 230 (24.5) | 310 (25.1) | 2148 (27.9) | 2469 (29.6) |
| 80+ | 1860 (10.2) | 129 (13.7) | 126 (10.2) | 904 (11.7) | 701 (8.4) |
| **Admission category, n (%)** | | | | | |
| Elective surgical | 7397 (40.6) | 0 (0.0) | 0 (0.0) | 2866 (37.2) | 4531 (54.3) |
| Emergency surgical | 1741 (9.6) | 197 (21.0) | 144 (11.7) | 856 (11.1) | 544 (6.5) |
| Emergency medical | 9083 (49.8) | 742 (79.0) | 1089 (88.3) | 3975 (51.6) | 3277 (39.2) |
| **Specialty, n (%)** | | | | | |
| Cardiothoracic surgery | 7369 [illegible] | 36 (3.8) | 48 (3.9) | 2469 [illegible] | 4816 [illegible] |
| Cardiology | 1300 (7.1) | 107 (11.4) | 70 (5.7) | 897 (11.7) | 226 (2.7) |
| Neurosurgery | 1089 (6.0) | 33 (3.5) | 74 (6.0) | 544 (7.1) | 438 (5.2) |
| Gastrointestinal surgery | 942 (5.2) | 101 (10.8) | 96 (7.8) | 497 (6.5) | 248 (3.0) |
| General Internal Medicine | 921 (5.1) | 159 (16.9) | 187 (15.2) | 354 (4.6) | 221 (2.6) |
| Vascular surgery | 897 (4.9) | 35 (3.7) | 32 (2.6) | 351 (4.6) | 479 (5.7) |
| Trauma | 774 (4.2) | 64 (6.8) | 76 (6.2) | 392 (5.1) | 242 (2.9) |
| Lung disease/surgery | 637 (3.5) | 61 (6.5) | 124 (10.1) | 298 (3.9) | 154 (1.8) |
| Neurology | 568 (3.1) | 38 (4.0) | 73 (5.9) | 266 (3.5) | 191 (2.3) |
| Haematology | 233 (1.3) | 42 (4.5) | 86 (7.0) | 87 (1.1) | 18 (0.2) |
| Gynaecology | 133 (0.7) | 9 (1.0) | 9 (0.7) | 67 (0.9) | 48 (0.6) |
| Urology | 132 (0.7) | 9 (1.0) | 16 (1.3) | 79 (1.0) | 28 (0.3) |
| Other medical specialty | 1286 (7.1) | 78 (8.3) | 139 (11.3) | 451 (5.9) | 618 (7.4) |
| Other surgical specialty | 331 (1.8) | 19 (2.0) | 26 (2.1) | 143 (1.9) | 143 (1.7) |
| From other ICU | 1121 (6.2) | 103 (11.0) | 112 (9.1) | 582 (7.6) | 324 (3.9) |
| **First 24hr physiology, median (IQR)** | | | | | |
| Maximum heart rate | 102 (89-119) | 127 (109-145) | 119 (103-135) | 103 (89-121) | 97 (87-110) |

| | | | | | |
|---|---|---|---|---|---|
| Minimum mean arterial pressure, mmHg | 56 (46-63) | 42 (33-52) | 50 (39-58) | 55 (45-62) | 58 (50-65) |
| Maximum FiO2 | 0.50 (0.41-0.61) | 0.70 (0.51-0.90) | 0.60 (0.41-0.90) | 0.51 (0.41-0.70) | 0.46 (0.40-0.55) |
| Minimum SpO2 | 0.94 (0.73-0.97) | 0.85 (0.72-0.94) | 0.94 (0.89-0.96) | 0.94 (0.73-0.97) | 0.95 (0.71-0.97) |
| Minimum PaO2, mmHg | 79 (68-95) | 69 (59-80) | 72 (63-86) | 78 (66-94) | 82 (71-99) |
| Minimum PaO2:FiO2 ratio | 195 (135-263) | 122 (85-183) | 158 (100-223) | 184 (126-251) | 215 (162-283) |
| Minimum GCS | 15 (10-15) | 13.0 (3.0-15.0) | 14.0 (7.0-15.0) | 15.0 (6.0-15.0) | 15 (14-15) |
| Maximum creatinine, µmol/L | 90 (72-119) | 131 (90-208) | 88 (65-135) | 92 (72-127) | 87 (72-106) |
| Minimum platelets | 150 (105-211) | 163 (93-237) | 210 (139-299) | 148 (100-213) | 146 (108-195) |
| Maximum bilirubin, µmol/L | 10.0 (6.0-17.0) | 12.0 (7.0-23.0) | 9.0 (5.0-16.0) | 11.0 (7.0-18.0) | 9.0 (6.0-14.0) |
| Maximum SOFA score | 6.0 (5.0-9.0) | 11.0 (8.0-13.0) | 7.0 (5.0-9.0) | 7.0 (5.0-10.0) | 5.0 (4.0-7.0) |
| Use of vasopressors, n (%) | 12254 (67.3) | 939 (100.0) | 679 (55.1) | 5442 (70.7) | 5194 (62.2) |
| Mechanical ventilation, n (%) | 15973 (87.7) | 858 (91.4) | 974 (79.0) | 6933 (90.1) | 7208 (86.3) |
| **Outcomes** | | | | | |
| Antibiotic escalation, first 24hr, n (%) | 5053 (27.7) | 939 (100.0) | 1233 (100.0) | 2881 (37.4) | 0 (0.0) |
| IV antibiotics for first 4d (*), n (%) | 2453 (13.5) | 582 (62.0) | 738 (59.9) | 1096 (14.2) | 37 (0.4) |
| IV antibiotics for at least 4d (**), n (%) | 5294 (29.1) | 795 (84.7) | 1041 (84.4) | 3077 (40.0) | 381 (4.6) |
| ICU length of stay, h, median (IQR) | 31 (22-114) | 154 (58-336) | 130 (55-282) | 57 (24-182) | 23 (20-41) |
| ICU mortality, n (%) | 2270 [illegible] | 358 (38.1) | 187 (15.2) | 1274 [illegible] | 451 (5.4) |

**Table S5.** Sensitivity analysis: Summary characteristics of ICU admissions by infection status when the criteria for suspected infection required both microbial cultures to be taken and the prescription of intravenous, non-prophylactic antibiotics. *IV (non-prophylactic) antibiotics consecutively for the first 4 days and admission length of stay >=4 days. †IV (non-prophylactic) antibiotics for at least 4 days in total or until ICU death/discharge.

| | **Sepsis status on admission (compared to *No sepsis*)** | **ICU mortality hazard ratio** *(95% CI)* | ***p* value** |
|---|---|---|---|
| a) All ICU admissions | Sepsis without shock | 0.69 *(0.61-0.77)* | < 0.001 |
| | Septic shock | 1.67 *(1.51, 1.84)* | < 0.001 |
| b) Emergency ICU admissions | Sepsis without shock | 0.46 *(0.41-0.52)* | < 0.001 |
| | Septic shock | 1.13 *(1.02, 1.25)* | 0.019 |

**Table S6.** Cause-specific Cox proportional hazard models predicting ICU mortality for *Sepsis without shock* and *Septic shock* compared to *No sepsis* among a) *all* (emergency & elective) ICU admissions and b) *emergency-only* ICU admissions, adjusted for age and sex.

| Admission SOFA score | ICU mortality hazard ratio *(95% CI)* | *p* value |
|---|---|---|
| *a) All admissions* | | |
| Max SOFA 4-7 | 1.41 *(1.05, 1.91)* | 0.024 |
| Max SOFA 8-11 | 2.85 *(2.13, 3.82)* | < 0.001 |
| Max SOFA 12-15 | 5.32 *(3.96, 7.16)* | < 0.001 |
| Max SOFA >15 | 10.15 *(7.39, 13.95)* | < 0.001 |
| *b) Septic shock** | | |
| Max SOFA 4-7 | n/a | n/a |
| Max SOFA 8-11 | 1.23 *(0.89, 1.71)* | 0.372 |
| Max SOFA 12-15 | 2.11 (*1.53-2.92*) | < 0.001 |
| Max SOFA >15 | 3.80 *(2.67-5.41)* | < 0.001 |
| *c) Sepsis without shock* | | |
| Max SOFA 4-7 | 1.04 *(0.61-1.76)* | 0.895 |
| Max SOFA 8-11 | 1.39 *(0.83-2.33)* | 0.199 |
| Max SOFA 12-15 | 1.81 *(1.04-3.16)* | 0.039 |
| Max SOFA >15 | 1.88 *(0.85-4.17*) | 0.117 |
| *d) Antibiotics without sepsis* | | |
| Max SOFA 4-7 | 1.92 *(0.98-3.76)* | 0.059 |
| Max SOFA 8-11 | 3.03 *(1.56-5.88)* | 0.001 |
| Max SOFA 12-15 | 5.95 *(3.04-11.60)* | < 0.001 |
| Max SOFA >15 | 13.58 *(6.68-27.60)* | < 0.001 |
| *e) No antibiotics* | | |
| Max SOFA 4-7 | 1.26 *(0.80-1.99)* | 0.322 |
| Max SOFA 8-11 | 5.58 *(3.58-8.70)* | < 0.001 |
| Max SOFA 12-15 | 14.61 *(9.18-23.3)* | < 0.001 |
| Max SOFA >15 | 47.58 *(26.6-85.2)* | < 0.001 |

**Table S7.** Cause-specific Cox proportional hazard models, adjusted for age and sex, predicting ICU mortality based on admission SOFA score (compared to baseline Max SOFA 0-3), both for all admissions (a) and grouped by sepsis status at admission (b-e). *Due to insufficient data in the '*Max SOFA 0-3'* group for patients with septic shock on admission, hazard models were calculated comparing to the baseline hazard of the '*Max SOFA 4-7*' group; all other hazard models in S7a-S7e were calculated in comparison to the baseline hazard of the '*Max SOFA 0-3*' group.

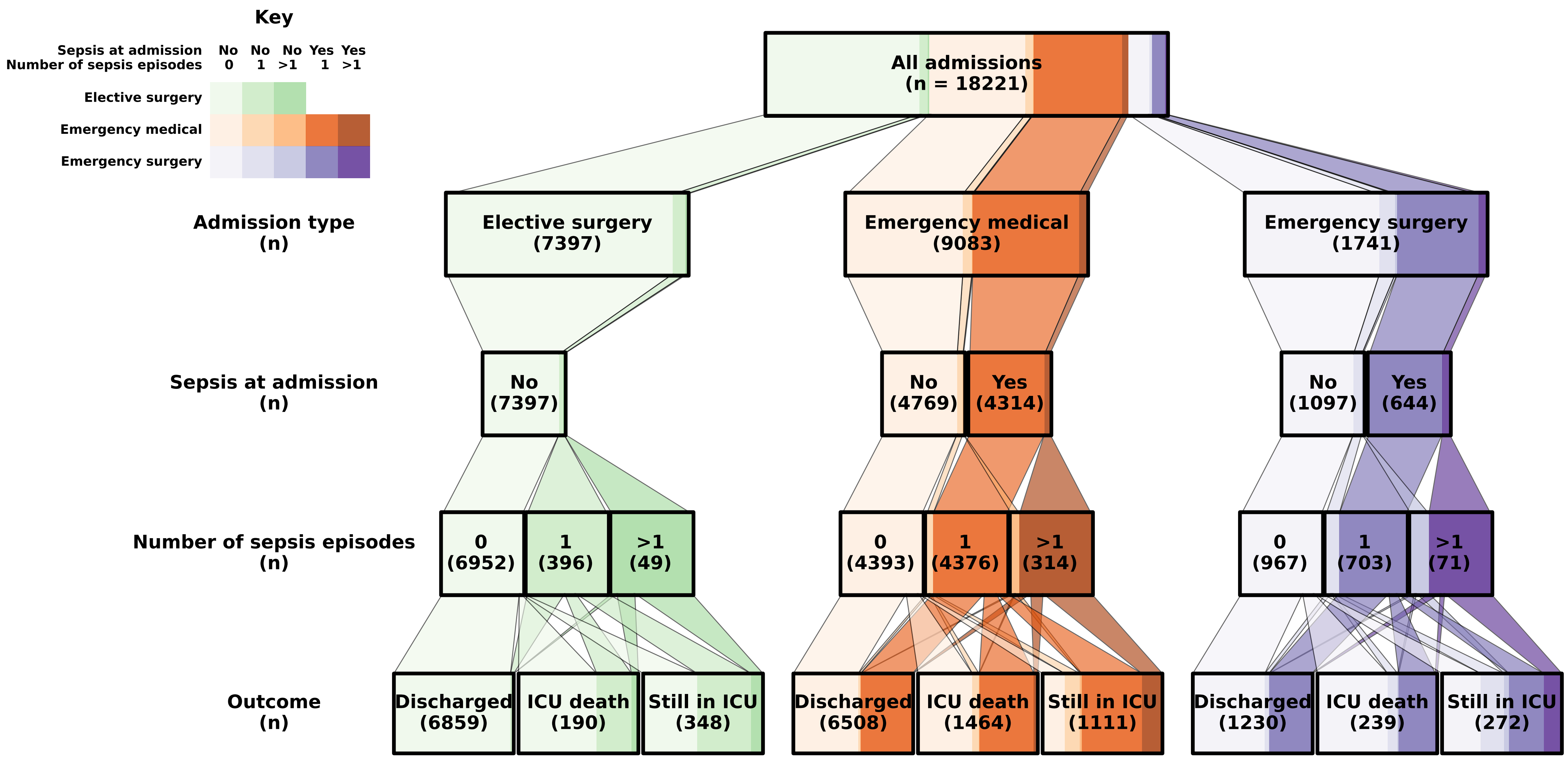

Key
Sepsis at admission
Number of sepsis episodes
No 0
No 1
No >1
Yes 1
Yes >1
Elective surgery
Emergency medical
Emergency surgery
All admissions (n = 18221)
Admission type (n)
Elective surgery (7397)
Emergency medical (9083)
Emergency surgery (1741)
Sepsis at admission (n)
No (7397)
No (4769)
Yes (4314)
No (1097)
Yes (644)
Number of sepsis episodes (n)
0 (6952)
1 (396)
>1 (49)
0 (4393)
1 (4376)
>1 (314)
0 (967)
1 (703)
>1 (71)
Outcome (n)
Discharged (6859)
ICU death (190)
Still in ICU (348)
Discharged (6508)
ICU death (1464)
Still in ICU (1111)
Discharged (1230)
ICU death (239)
Still in ICU (272)
